# Prefrontal suppression in short-video viewing: unraveling the neural correlates of self-control

**DOI:** 10.1101/2023.10.30.23296738

**Authors:** Conghui Su, Binyu Teng, Hui Zhou, Fengji Geng, Yuzheng Hu

**Affiliations:** Department of Psychology and Behavioral Sciences, Zhejiang University, Hangzhou 310058, China; Department of Curriculum and Learning Sciences, College of Education, Zhejiang University, Hangzhou 310007, China; The State Key Lab of Brain-Machine Intelligence, College of Education, Zhejiang University, Hangzhou 310007, China; MOE Frontiers Science Center for Brain Science & Brain-Machine Integration, Zhejiang University, Hangzhou 310058, China; Brain and Cognitive Research Institute, School of Medicine, Hangzhou City University, Hangzhou 310015, China

**Keywords:** short-video watching, self-control, amygdala, dorsolateral prefrontal cortex, mindfulness

## Abstract

The recent surge in short-video application usage has raised concerns about potential mental health risks. Using a novel video-watching task, we investigated the neuropsychological mechanisms underlying self-control during short-video viewing from a dual-system perspective. Results revealed watching preferred videos significantly activated the amygdala (System I) and deactivated the control regions (System II), with individuals with lower trait self-control being suppressed more. Dynamic causal modelling revealed the amygdala inhibited control regions during preferred viewing, while control regions downregulated the amygdala during less-preferred viewing. The control regions also demonstrated enhanced activation during cognitive control and inner-state monitoring tasks, with the latter correlating with trait self-control. These findings suggest preference-based video-watching suppresses prefrontal areas that represent rules and support self-awareness, enabling bottom-up limbic processes to dominate attention. This study provides insights into the neuropsychological impacts of short-video applications use, informing policies and interventions to promote healthier technology use and mitigate potential adverse effects.

## Main

As short-video applications continue to surge in popularity, concerns about the consequences of excessive screen time are growing ^1^. Globally, short-video platforms, such as YouTube Shorts, TikTok, and Instagram Reels, have been utilized by billions of users. In China alone, there are about one billion online short-video watchers, accounting for 94.8% of its internet users ^2^. In addition to growing in user population, the average time spent with these apps is also on the rise. For example, it has been reported that an average user opens TikTok 19 times and spends an average of 95 minutes on the app in a single day ^3^. These applications tap into the human desire for novelty and immediate gratification, making it difficult for individuals to resist the allure of binge-watching. Additionally, the algorithms employed by these platforms are designed to personalize and optimize the content based on users’ preferences, further reinforcing the cycle of engagement ^4^. That is, the constant availability of engaging content, combined with the accurate recommendation strategies employed by these platforms, may erode self-control, manifesting in prolonged use or addictive-like binge-watching in some vulnerable individuals ^5,6^. Studies have shown that excessive short-video watching can negatively affect mental health ^7,8^ and cognitive functions ^9,10^. This emerging public health concern underscores an urgent need to understand the underlying neuropsychological mechanism of self-control during short-video watching. Such knowledge could inform the development of strategies to minimize harmful usage patterns, promote healthier use behavior, and guide policy-making around digital media consumption.

Excessive using behaviors, ranging from over-use of Internet to severe drug addiction, are often linked to the psychological construct of self-control ^11,12^, an important ability that allows individuals to regulate their thoughts, emotions, and behaviors for the pursuit of long-term goals ^13^. As an umbrella construct bridging concepts from different research fields, questionnaires are widely used to characterize this competence with different names (e.g., impulsivity, conscientiousness, self-regulation, willpower, et. al) ^14,15^. High levels of self-control are generally associated with positive outcomes in various aspects of life, contributing to success and overall well-being ^16–18^. Conversely, individuals with lower levels of self-control are more prone to develop addictive behaviors ^11,19^. For example, previous studies have shown that adolescents with higher trait self-control are less likely to develop dependence on short-videos ^6^ and are more capable of mitigating the negative impacts of their environment ^20^. Although self-reported self-control serves as a reliable predictor of real-life outcomes ^15–17,21^, the underlying neuropsychological mechanisms of self-control remains incomplete to explain how modern technology such as short-video apps grab and hold large amount of attention.

Among many theoretical frameworks that have sought to shed light on the mechanisms underlying self-control ^22^, the dual-system model ^23^ has gained significant recognition. Despite its variations and revisions, the model’s core premise remains the same — self-control is driven by two distinct systems known as System I and System II ^22^. System I, also referred to as the automatic system, is highly responsive to environmental stimuli and gives rise to habitual behaviors and urges to seek immediate gratification ^22^. Neuroimaging studies have provided evidence linking this system to subcortical regions involved in emotion and reward processing, particularly the amygdala and ventral striatum ^24^. On the other hand, System II, the reflective system, is primarily guided by goals or rules stored in working memory ^25^. It plays a crucial role in flexible responses, such as monitoring and resolving conflicts, overriding habitual tendencies and resisting temptations, with the prefrontal cortex and cingulate cortex as the main neural correlates ^26^.

Self-control involves a balance between the two systems ^27^, and it has been commonly thought that the exertion of top-down control by System II over System I is imperative for successful self-control^24^. By leveraging the cognitive abilities associated with System II, individuals can effectively regulate their behaviors ^28^ and resist the pull of immediate gratification ^29^. Therefore, previous research has underscored the System II’s pivot role in self-control, examining its activity in resolving conflicts or inhibiting distractions through laboratory cognitive tasks, such as Flanker task, Go/No-Go task, and Stop Signal task ^30^. However, self-control measured with these laboratory tasks has shown poor correlation with trait self-control assessed using questionnaires ^31–33^, calling a necessity of examining the dynamic interaction between the two systems and its implications for real-life behaviors by using new task paradigms with better ecological validity ^34^.

To this end, we proposed a new perspective that emphasized the interplay between the two systems in regulation/dysregulation of excessive using behavior. In the case of short-video watching, a user either watches a clip to its end or selects an alternative, depending on the degree of affective satisfaction at the moment. When facing preferred audio-visual stimuli, the System I would rapidly activate, and a resultant bottom-up process would prevent System II from disrupting the viewing process. Instead, less satisfied content would trigger a top-down process from System II to make an adjustment to the viewing process. Through such a dynamic interplay between the two systems, the use of short-video apps can swiftly modulate brain activity to create a satisfied psychological state. While short duration of videos allows quick iterations between stay and switch to modulate the brain, tremendous variety, and an endless list personalized by artificial intelligent (AI) recommendation within short-video apps can significantly accelerate this process and enhance engagement.

To depict the activation patterns and between-system interplay described above, we designed a novel and naturalistic video-watching task that allowed participants to voluntarily select their preferred content. By mimicking daily use behavior, this paradigm would recapitulate brain activity patterns emerging during video viewing in real-life situation. Given that losing track of one’s agenda and surroundings is a typical phenomenon during short-video watching, the awareness of one’s own behavior and the representation of one’s long-term goals/plans would be weakened or overridden. As these functions are critical for self-control, it is reasonable to hypothesize that brain activation during this video task would manifest one’s trait self-control level. In addition, as these functions are supported by System II ^35^, the activation of System II may be suppressed when engrossed in video watching. To contrast the brain activation of the two systems during video watching to that when performing cognitive tasks, we incorporated two traditional cognitive tasks, the Go/No-Go task and the Dots task (rule-switching). In addition, the ability to monitor one’s present state is a prerequisite for adjusting ongoing thoughts or actions ^36^. Therefore, it is plausible to hypothesize that brain regions in System II suppressed during video watching are the neural substrates of awareness of one’s inner-state. To test this hypothesis, we conducted a heartbeat detection task requiring the awareness of one’s inner-state. We recorded participants’ brain activity during these tasks using functional magnetic resonance imaging (fMRI). An overall study procedure is illustrated in **Figure 1**.

**Figure 1.**
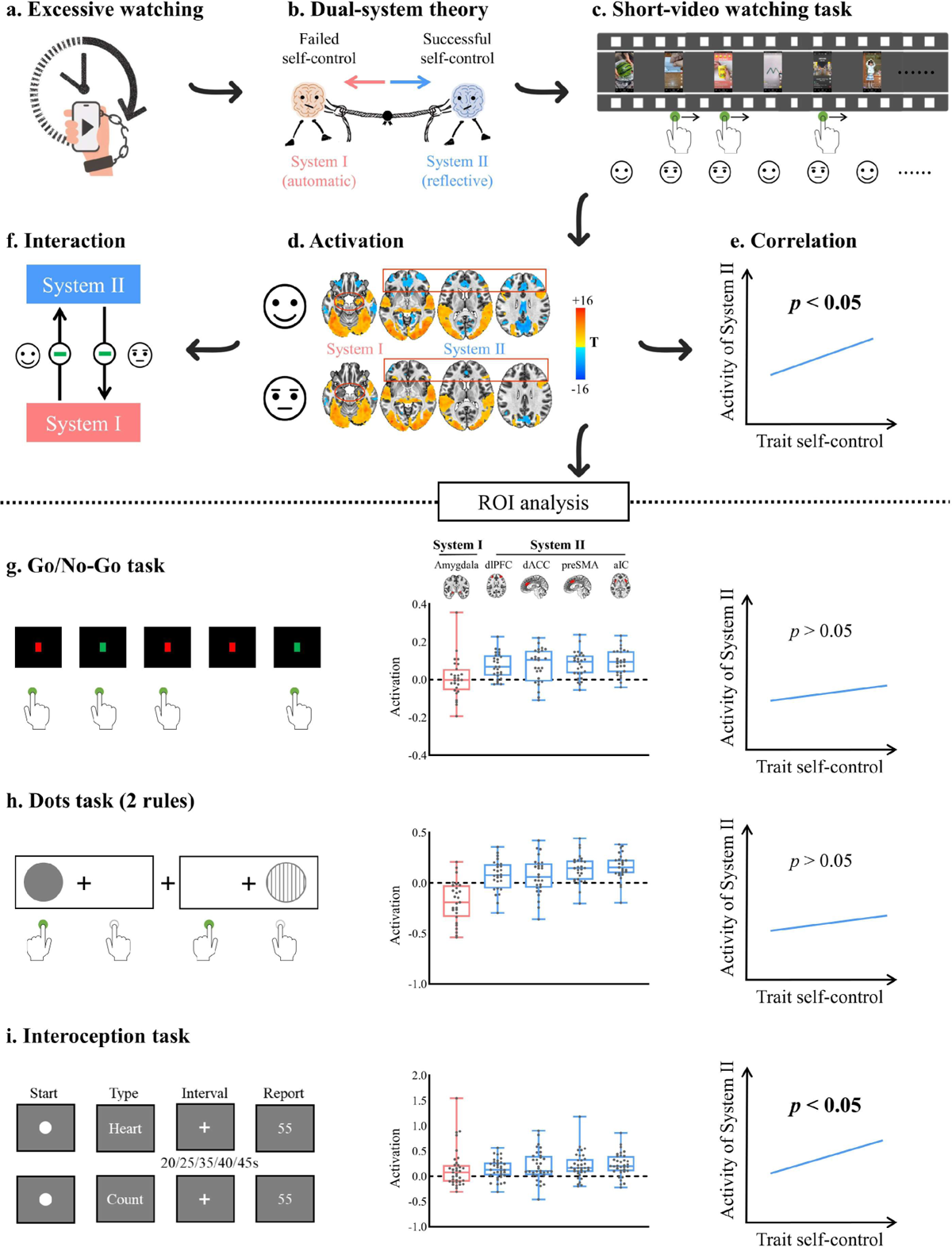
Analysis Flowchart and Result Schematics. (a) The lack of self-control over short-video watching is a public concern. (b) The dual-system model attributes self-control to the balance between the automatic (System I) and reflective (System II) systems. (c) A novel video-watching task was designed to characterize brain activity during the viewing of preferred and less-preferred content. (d) General linear modelling revealed that video-watching activated System I (marked with red circles) while deactivating System II (marked with red rectangles). (e) The brain activity of System II regions during watching short-videos correlated with trait self-control. (f) Dynamic causal modelling revealed that in the face of preferred content, System I inhibited System II. Conversely, when facing less-preferred content, System II down-regulated System I. (g) Regions of interest (ROIs) analysis for the Go/No-Go task showed significant activation of System II during the response inhibition process, but no correlation with self-control. (h) The Dots task, demanding cognitive flexibility to switch between two rules, deactivated System I and activated System II, but showed no correlation with trait self-control. (i) An interoception task requiring inner-state awareness of heartbeat activated both systems, and the System II activity during heartbeat perception significantly correlated with trait self-control.

Our analyses of the fMRI data from the video-watching task demonstrated that the amygdala, a key region involved in emotional processing within System I, showed significant activation, particularly when participants were watching videos they liked. In contrast, the control regions within System II, including the dorsolateral prefrontal cortex (dlPFC), dorsal anterior cingulate cortex (dACC), anterior insular cortex (aIC), and pre-supplementary motor area (preSMA), exhibited deactivation. Trait self-control measures were found to correlate with brain activities of these four regions, but not of the amygdala. In contrast to the video task, the two cognitive tasks significantly activated the four control regions under the conditions involving inhibitory control, conflict resolution, or rule-switching. However, no significant correlation between trait self-control and brain activation in abovementioned control regions under these conditions was found. Next, to better depict the interplay between System I and System II during video watching, we applied dynamic causal modelling (DCM) ^37^ to characterize the effective connectivity between these regions. Our DCM analysis revealed that the amygdala exerted an inhibitory influence on all the four control regions during participant’s viewing of their preferred videos. In contrast, when participants were viewing less preferred videos, the dlPFC and dACC down-regulated the amygdala’s activation. Lastly, to test our hypothesis that System II suppressed by video-watching task support awareness of one’s inner-state, we compared the neural responses in the heartbeat detection condition and the control number-counting condition, and found higher activity of System II in the heartbeat detection condition. Further, the activation in System II during heartbeat perception positively and significantly correlated with levels of trait self-control.

In summary, our results shed light on the neural mechanisms behind the over-viewing behavior of short videos from the perspective of interaction between the two systems of self-control, and provide valuable insights to promote healthier viewing behaviors.

## Results

### Different measures of trait self-control were significantly correlated

Trait self-control is a multifaceted construct that can be measured in various ways. In our study, we used three well-established questionnaires to measure different aspects of trait self-control, including general self-control, impulsiveness, and mindfulness. Our results showed significant correlations between these different measures. Specifically, general self-control was negatively correlated with impulsiveness (*r* = −0.692, *p* < 0.001) and positively correlated with mindfulness (*r* = 0.493, *p* = 0.009), with the latter two negatively correlating with each other (*r* = −0.625 *p* < 0.001). These correlations remained significant after controlling for age, gender, anxiety, and depression, except for the correlation between general self-control and mindfulness (*r* = 0.370, *p* = 0.082).

### Weak relations between trait self-control and behavioral performances of laboratory cognitive tasks

Motivated by the traditional assumption in the literature that individuals with higher trait self-control might perform better on tasks requiring high cognitive control, we also explored how self-reported self-control related to cognitive control abilities measured using laboratory tasks. We used the Go/No-Go task, which measures the ability to inhibit prepotent motor responses (inhibitory control), and the Dots task, which assesses the ability to monitor and resolve conflict (conflict monitoring and resolution) and switch between different task rules (cognitive flexibility). While previous findings on the correlation between trait self-control and cognitive control task performance are mixed, we found the relationships between trait self-control and cognitive control measures with relatively small effect size. Specifically, only the cognitive control measured by the mean response times in the Mixed condition of the Dots task was significantly related to general self-control (*r* = 0.424, *p* = 0.044) and impulsiveness (*r* = −0.455, *p* = 0.029). The other correlations were not significant (*p* > 0.05). Detailed information about performance in the cognitive control tasks and their relationships with trait self-control is presented in Table S2.

### Voxel-wise analyses showed the activation of amygdala in System I and the deactivation of prefrontal cortex in System II during watching short-videos

Next, we sought to investigate the associations between trait self-control and neural activities from a dual-system perspective. We designed a naturalistic short-video watching task that allowed participants to choose which videos to watch based on their personal preferences, without any external rules constraining their choices. The videos were categorized into Like and Dislike groups according to each participant’s choices. Videos that participants watched from beginning to end were categorized as Like videos (average portion = 35.2%), while those that participants watched less than 50% of were classified as Dislike videos (average portion = 46.2%).

Using voxel-wise general linear modelling, we examined the brain activity associated with both Like and Dislike conditions and compared their activation patterns (Figure 2, corrected *p* < 0.05). In both conditions, visual cortex, auditory cortex, and middle temporal lobe were activated, whereas posterior cingulate cortex, precuneus, inferior parietal cortex, and cerebellum were deactivated (Figure 2a and 2b, one-sample t-test).

**Figure 2.**
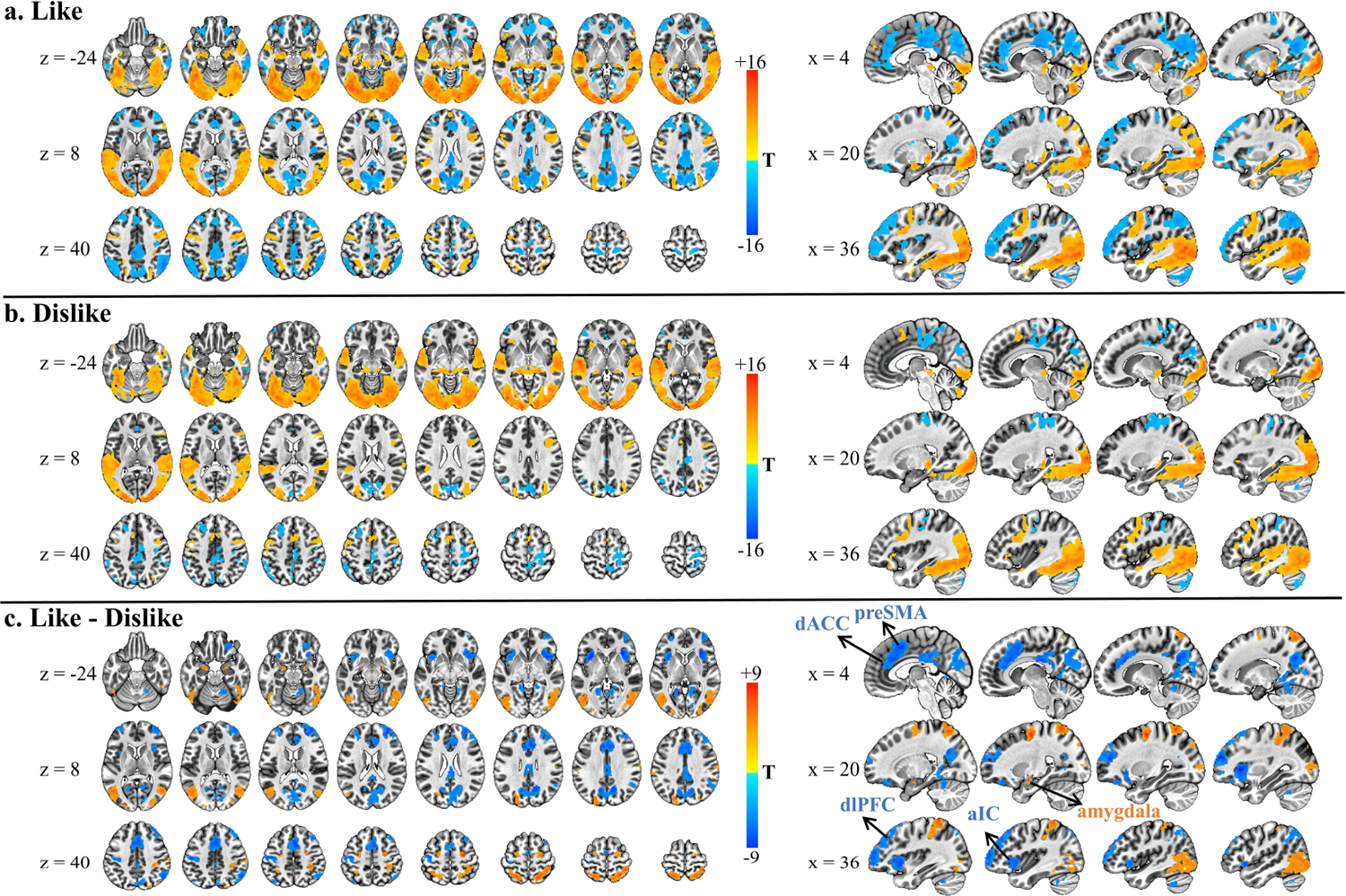
Activation Maps of Group-level Analysis in the Video-watching Task (*p* < 0.05 corrected). (a) Activation for Like relative to fixation baseline. (b) Activation for Dislike relative to fixation baseline. (c) Contrast of “Like > Dislike”. As illustrated in (c), the amygdala, a key region of automatic System I, displayed heightened activation whereas prefrontal regions including dACC, preSMA, dlPFC and aIC, in the reflective System II showed pronounced suppression when viewing preferred video content (i.e., in the Like condition). Abbreviations: dACC, dorsal anterior cingulate cortex; preSMA, pre-supplementary motor area; dlPFC, dorsolateral prefrontal cortex; aIC, anterior insular cortex.

A paired-sample t-test was used to assess the statistical differences between the two conditions. Results showed that, amygdala (a key region in System I), middle temporal cortex, and dorsal attention network (intraparietal sulcus and frontal eye fields) were activated in a higher extent in the Like condition (Figure 2c). Contrarily, frontal regions in System II, including dlPFC (BA 9,46), dACC (BA 32,24), preSMA (BA 6), and aIC showed significant differential activation between the Like and Dislike conditions. Specifically, these regions were deactivated in the Like condition, but such deactivation was less pronounced in the Dislike condition, with preSMA and aIC even exhibiting positive activation (Figure 2c). Expect for these frontal regions, posterior cingulate cortex and precuneus also showed similar between-condition difference (see supplementary Table S1).

### ROI-wise analyses revealed the deactivation of System I and the activation of System II during cognitive control tasks

In the video-watching task, participants freely selected videos based on personal preference. In contrast, traditional cognitive tasks require rule-based information processing. To contrast neural activation patterns with traditional cognitive tasks, we focused on five regions of interest (ROIs). The amygdala, a central region of System I, and four key regions of System II—dlPFC, aIC, dACC, and preSMA—were selected (Figure 3, for a more detailed justification, please refer to *Methods* and supplementary Figure S3). We then extracted brain activation data from each region for each condition in the Go/No-Go and Dots tasks for statistical analysis.

**Figure 3.**
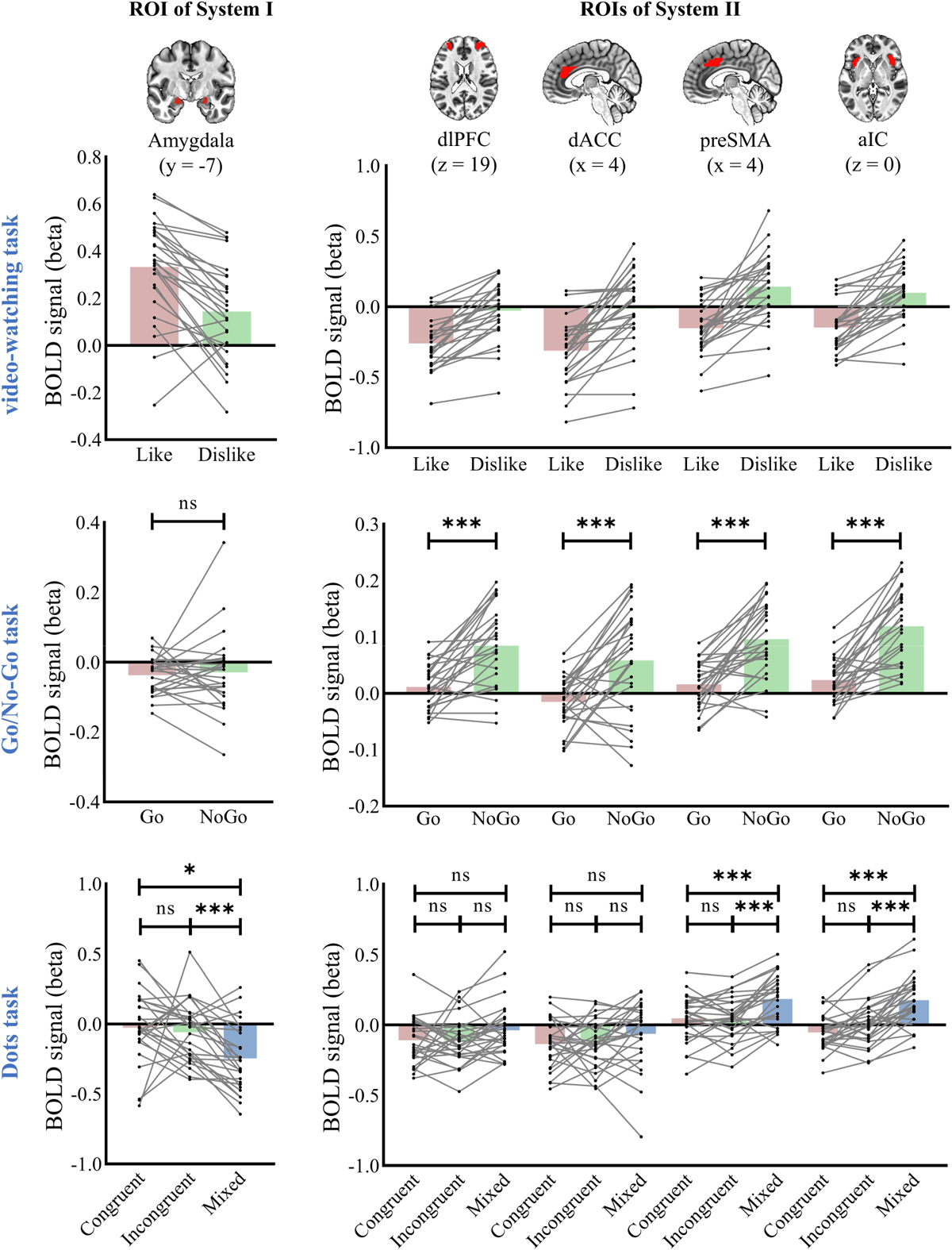
Activation Patterns of the Two Systems Across Three Tasks. Amygdala from System Ⅰ (the left column), and other four ROIs from System Ⅱ (the right column) including dorsolateral prefrontal cortex (dlPFC), dorsal anterior cingulate cortex (dACC), pre-supplementary motor cortex (preSMA) and anterior insular cortex (aIC), were identified and visualized in MNI space. During watching short-videos (the top row), System Ⅰ was activated under both Like and Dislike conditions, with higher activation in the former. Conversely, System Ⅱ was deactivated under the Like condition but partly activated under the Dislike condition. In the Go/No-Go task (the middle row), System Ⅰ displayed similar deactivations in both Go and NoGo trials, and System Ⅱ showed higher activation in NoGo trials requiring response inhibition. In the Dots task (the bottom row), the Mixed condition demanding on cognitive flexibility deactivated System Ⅰ, while activating the preSMA and aIC within System Ⅱ. ns, nonsignificant; * *p* < 0.05, ** *p* < 0.01, *** *p* < 0.001.

Contrasting to its positive activation during video watching, the amygdala was predominantly suppressed, particularly in the Mixed condition of the Dots task, where rule-switching was required. This distinction highlights the amygdala’s varied roles in self-directed video-watching versus rule-driven cognitive tasks.

For the regions in System II, two distinct activation patterns emerged across various task conditions. Specifically, dlPFC and dACC showed one pattern: deactivation during the viewing of Like videos (dlPFC: *t* = −7.69, *p* < 0.001, Cohen’s d = 1.48; dACC: *t* = −6.95, *p* < 0.001, Cohen’s d = 1.34), near baseline activation when viewing Dislike videos (dlPFC: *t* = −0.74, *p* = 0.48, Cohen’s d = 0.14; dACC: *t* = −0.29, *p* = 0.78, Cohen’s d = 0.19), positive activation in the NoGo condition of the Go/No-Go task (dlPFC: *t* = 6.09, *p* < 0.001, Cohen’s d = 1.17; dACC: *t* = 3.28, *p* = 0.003, Cohen’s d = 0.63), and a non-significant trend of deactivation in the Dots task. This pattern suggests that dlPFC and dACC suppressed during preferred video viewing are the neural substrates for rule-guided response inhibition.

In contrast, the preSMA and aIC exhibited another pattern. These two regions were significantly deactivated when viewing Like videos (preSMA: *t* = −4.14, *p* < 0.001, Cohen’s d = 0.80; aIC: *t* = −4.42, *p* < 0.001, Cohen’s d = 0.85) and significantly activated when viewing Dislike videos (preSMA: *t* = 2.99, *p* = 0.006, Cohen’s d = 0.58; aIC: *t* = 2.58, *p* = 0.016, Cohen’s d = 0.50). In the Go/No-Go task, both regions demonstrated positive activation in both Go and NoGo conditions, with a more pronounced activation in the NoGo condition (preSMA: *t* = 5.85, *p* < 0.001, Cohen’s d = 1.13; aIC: *t* = 7.31, *p* < 0.001, Cohen’s d = 1.41). In the Dots task, these regions showed significantly higher activation in the Mixed condition, compared to both Congruent and Incongruent conditions (preSMA: *F* = 16.19, *p* < 0.001, partial η^2^= 0.38; aIC: *F* = 35.37, *p* < 0.001, partial η^2^= 0.58).

Notably, despite the positive activation in the Go condition of the Go/No-Go task, these regions displayed neither activation nor deactivation in the Congruent (preSMA: *t* = 1.53, *p* =0.14, Cohen’s d = 0.29; aIC: *t* = −2.24, *p* = 0.034, Cohen’s d = 0.43) and Incongruent conditions (preSMA: *t* = 1.56, *p* = 0.13, Cohen’s d = 0.30; aIC: *t* = 0.47, *p* = 0.64, Cohen’s d = 0.09) of the Dots task. This pattern implies that the preSMA and aIC, when suppressed during preferred video viewing, play an important role in cognitive flexibility, particularly in transitioning to alternate response rules or affective states.

### Trait self-control was robustly related to the activity of System II during video task but not during Go/No-Go and Dots tasks

Given that self-control primarily pertains to one’s ability to voluntarily adjust their internal thoughts, emotions, and behaviors, we proceeded to investigate whether brain activation during a more naturalistic task, such as watching videos, would correlate more closely with trait self-control.

In the Dislike condition, the activities of the four ROIs in System II were significantly related to general self-control, mindfulness, and impulsiveness (**Table 1**). In the Like condition, while the dlPFC activation was moderately related to general self-control and impulsivity, activities of all the four regions were significantly and positively related to mindfulness (**Table 1**). However, no significant correlation was found between trait self-control and amygdala activity in either Like or Dislike conditions.

**Table 1.** Statistical information of neural activities of five ROIs from two systems across tasks.

| Tasks | Conditions | Regions | Activation (beta value) |  |  | Partial Correlation |  |  |
| --- | --- | --- | --- | --- | --- | --- | --- | --- |
|  |  |  | M | SD | T-test | BSCS | BIS-11 | FFMQ |
| Video-watching | Dislike | amygdala | 0.14 | 0.20 | 3.73 | -0.109 | 0.075 | 0.035 |
|  |  | dlPFC | -0.03 | 0.21 | -0.74 | 0.657 *** | -0.68 *** | 0.417 * |
|  |  | dACC | -0.02 | 0.27 | -0.29 | 0.539 ** | -0.565 ** | 0.398 |
|  |  | preSMA | 0.14 | 0.25 | 2.99 | 0.384 | -0.628 ** | 0.467 * |
|  |  | aIC | 0.10 | 0.20 | 2.58 | 0.55 ** | -0.671 *** | 0.51 * |
|  | Like | amygdala | 0.33 | 0.21 | 8.30 | -0.257 | -0.049 | 0.104 |
|  |  | dlPFC | -0.26 | 0.18 | -7.69 | 0.42 * | -0.521 * | 0.572 ** |
|  |  | dACC | -0.31 | 0.23 | -6.95 | 0.227 | -0.316 | 0.443 * |
|  |  | preSMA | -0.15 | 0.19 | -4.14 | 0.124 | -0.272 | 0.512 * |
|  |  | aIC | -0.15 | 0.17 | -4.42 | 0.282 | -0.334 | 0.544 ** |
| Go/No-Go | NoGo - Go | amygdala | 0.01 | 0.11 | 0.39 | -0.156 | 0.259 | -0.24 |
|  |  | dlPFC | 0.07 | 0.07 | 5.67 | 0.39 | -0.378 | 0.069 |
|  |  | dACC | 0.07 | 0.09 | 4.28 | 0.335 | -0.545 ** | 0.412 |
|  |  | preSMA | 0.08 | 0.07 | 5.85 | 0.386 | -0.272 | -0.119 |
|  |  | aIC | 0.10 | 0.07 | 7.31 | 0.195 | -0.368 | -0.162 |
| Dots | Incongruent - Congruent | amygdala | -0.03 | 0.33 | -0.51 | 0.057 | -0.141 | -0.196 |
|  |  | dlPFC | -0.001 | 0.19 | -0.04 | 0.078 | -0.16 | 0.044 |
|  |  | dACC | 0.005 | 0.20 | 0.12 | 0.286 | -0.229 | -0.079 |
|  |  | preSMA | -0.002 | 0.12 | -0.10 | 0.229 | -0.127 | 0.109 |
|  |  | aIC | 0.07 | 0.15 | 2.37 | -0.051 | -0.026 | 0.217 |
|  | Mixed - Incongruent | amygdala | -0.19 | 0.20 | -4.75 | -0.187 | 0.293 | 0.058 |
|  |  | dlPFC | 0.07 | 0.15 | 2.46 | -0.003 | 0.055 | -0.239 |
|  |  | dACC | 0.07 | 0.18 | 1.89 | -0.187 | 0.322 | -0.035 |
|  |  | preSMA | 0.14 | 0.14 | 5.12 | -0.037 | -0.11 | -0.052 |
|  |  | aIC | 0.16 | 0.13 | 6.45 | 0.178 | -0.024 | -0.172 |

Note: \* $p < 0.05$ , \*\* $p < 0.01$ , \*\*\* $p < 0.001$ . M, mean value; SD, standard deviation; BSCS, Brief Self Control Scale; BIS-11, Barratt Impulsiveness Scale; FFMQ, Five Factor Mindfulness Questionnaire. Age, gender, anxiety, and depression scores were used as covariates in the partial correlational analysis.

To test the specificity of video task in the brain-behavioral associations, we conducted a complementary analysis on the correlation between trait self-control and brain activation in the Go/No-Go and Dots tasks. Only the dACC activity in the Go/No-Go task showed a significant and negative correlation with impulsivity (*r* = −0.545, *p* = 0.007). Aside from this, we did not find any significant relationship between neural activities of two systems and general self-control, impulsivity, or mindfulness (**Table 1**).

To assess the robustness of the observed relationship between self-reported self-control and the neural activities of System II under the Dislike condition, we conducted a leave-one-out cross validation analysis. Our results suggest that regional activities of System II in the Dislike condition, with four additional variables (i.e., age, gender, anxiety, and depression) as covariates, can provide reliable prediction on trait self-control (supplementary Figure S1).

### The interaction between System I and System II during Like vs. Dislike video-viewing

To test the hypothesis that System I would suppress System II during video watching, we utilized DCM to elucidate how different video-watching states modulate the interaction between the two systems. DCM, a Bayesian-based technique, allows for the estimation of directional interactions (i.e., effective connectivity) between brain regions at a neurobiological level. We applied a full model (see supplementary Figure S2) to each participant’s data and performed a group-level Parametric Empirical Bayes (PEB) analysis. Parameters surpassing a posterior probability of 95% are depicted in Figure 4 and supplementary Figure S2.

**Figure 4.**
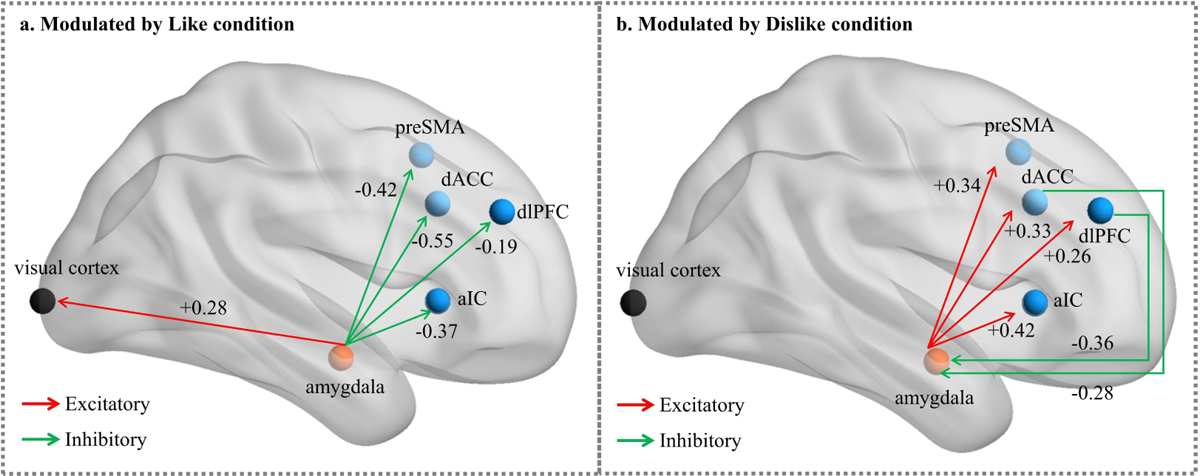
Modulatory effects of Like/Dislike Conditions on the Effective Connectivity Between System I and II, as Estimated by the PEB Group-level Matrix B. (a) Modulated by the Like condition, System I exerted inhibitory effect on System II. (b) Modulated by the Dislike condition, System I excited System II but also received inhibitory inputs from System II (primarily the dlPFC and dACC). Abbreviations: dlPFC, dorsolateral prefrontal cortex; dACC, dorsal anterior cingulate cortex; preSMA, pre-supplementary motor area; aIC, anterior insular cortex.

Our results delineate a reciprocal excitation/inhibition between amygdala (System I) and the four System II regions (i.e., dlPFC, dACC, preSMA, and aIC) under the Like and Dislike conditions. During engagement with preferred content in the Like condition, the amygdala, while showing excitatory connection with the visual cortex, imposed inhibitory effects on the System II regions to varying degrees (negative values in Figure 4a, in units of Hz). In contrast, when interacting with less preferred content in the Dislike condition, this dynamic was reversed: the amygdala exerted excitatory influences on the four System II regions while concurrently receiving inhibitory inputs from the dACC and dlPFC (negative values in Figure 4b, in units of Hz). In addition, the connectivity from aIC to preSMA was increased in the Dislike condition (supplementary Figure S2d). These patterns demonstrate a context-dependent excitatory and inhibitory interplay between the two systems, depicting a dynamic feature of self-control process in an ever-changing real-life environment.

### System II was enhanced in the interoception task and its neural activity was significantly correlated with trait self-control

Our hypothesis posits that certain brain regions, suppressed during video watching, support the awareness of one’s inner state, a crucial precursor to the exertion of self-control. To explore this, we conducted a heartbeat detection task that required participants to accurately perceive their own heartbeat during specified time intervals. This task effectively engaged participants’ internal monitoring systems. We also implemented a control condition that involved number counting, to separate the influence of mental counting when examining brain activation.

We first examined the four regions of System II, which were suppressed during the video-watching task. If these regions truly support the function of inner-state awareness, they should display increased activation in the heartbeat detection condition, and this activation should correlate positively with trait mindfulness and self-control. Consistent with this expectation, the ROI analysis revealed that all the four regions showed increased activity during the heartbeat detection compared to the control condition (dlPFC: *t* = 4.17, *p* < 0.001, Cohen’s d = 0.71; dACC: *t* = 4.14, *p* < 0.001, Cohen’s d = 0.7; preSMA: *t* = 4.74, *p* < 0.001, Cohen’s d = 0.8; aIC: *t* = 5.86, *p* < 0.001, Cohen’s d = 0.99; Figure 5). Importantly, these activations were positively and significantly correlated with general self-control and mindfulness (Figure 5, **Table 2**). Such an association was not observed in the number counting condition.

**Figure 5.**
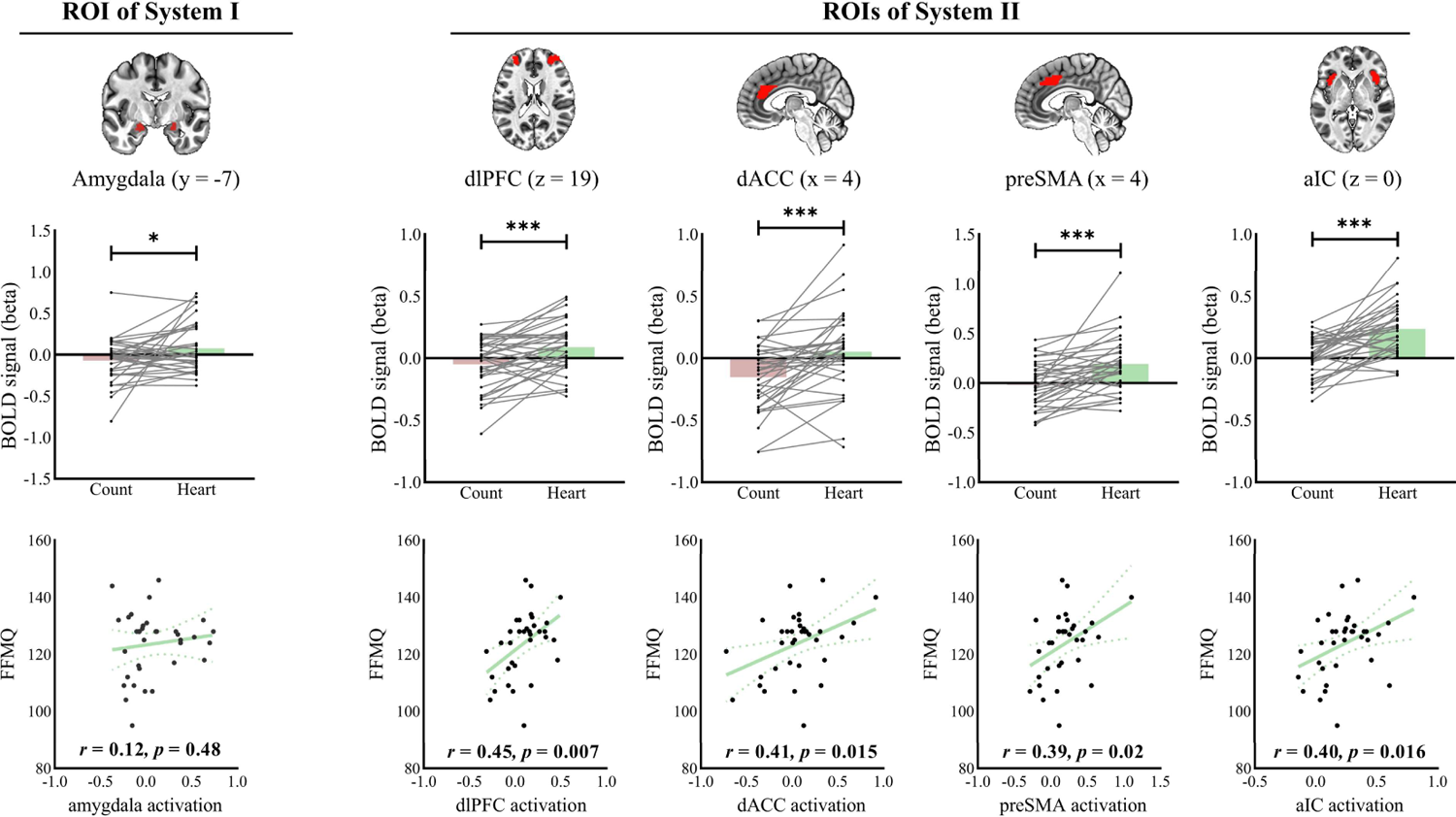
ROI-analysis for the Interoception Task. The top panel’s bar plots display the BOLD responses distribution of the two systems across conditions. Higher activations were observed in the heartbeat detection (Heart) condition compared to number counting (Count) condition. Only the neural activity in the four regions of System II significantly correlated with levels of trait mindfulness, as measured by the FFMQ score. Individual data points showing activations (beta values) of the two systems were provided in both bar and scatter plots.

**Table 2.**
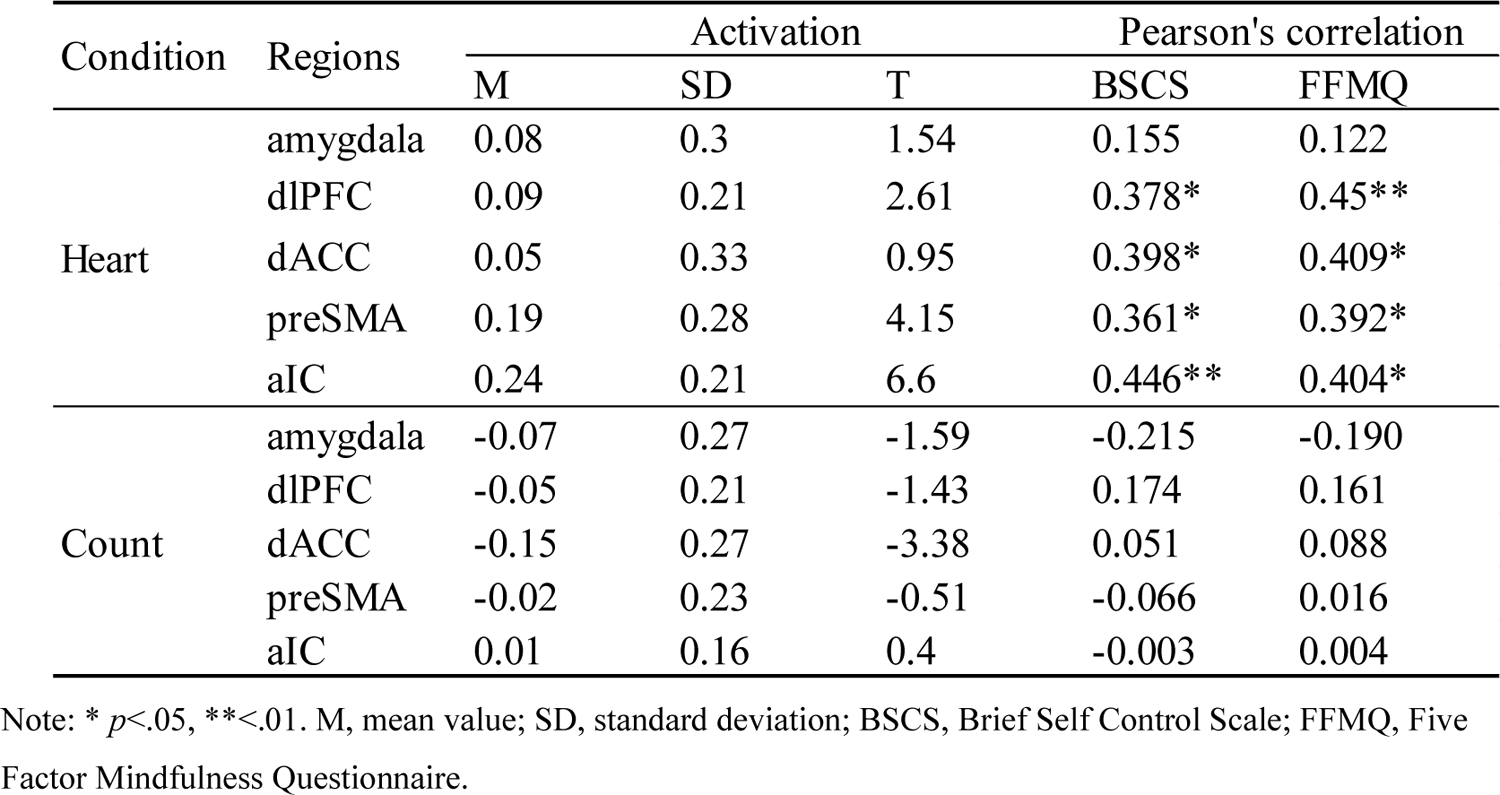
Correlations between trait self-control and brain activation in the interoception task.

We further probed the response of the amygdala in System I, given its significant activation and deactivation during video watching and traditional cognitive tasks, respectively. Interestingly, the amygdala maintained a moderate, but significantly higher, level of activation in the heartbeat detection condition compared to the control condition (*t* = 2.34, *p* = 0.03, Cohen’s d = 0.4). This suggests a unique state distinct from cognitive tasks. However, similar to the observations from the video-watching task, the amygdala activation did not correlate with general self-control or mindfulness (*p* > 0.05).

## Discussion

In present study, we found that when participants were viewing their preferred content, the amygdala (System I) was activated, while System II regions were deactivated. Conversely, when viewing less preferred content, the amygdala was less activated and System II regions were less deactivated. Our DCM results further revealed that during preferred content viewing, the amygdala exerted an inhibitory influence on the four regions of System II, and two of them (dlPFC and dACC) downregulated the amygdala when viewing less preferred content. In addition, the System II regions were activated to varying extent in two traditional cognitive tasks and an interoceptive task, but only the activation during the interoceptive task correlated with trait self-control. In below sections, we discussed the implications of these findings to better understand the worldwide rapid escalation of short-video usage, and we proposed a theoretical model from the dual-system perspective to account for the neuropsychological processes of video-watching behavior reinforced by AI recommendation.

### Implications of the activation of system I and deactivation of System II in understanding of video-watching behavior

Our findings suggest that the activation of System I, particularly the amygdala, is instrumental in shaping the engaging experience of video watching. This concurs with previous literature which emphasizes the role of the amygdala in emotional processing and motivation ^38^. The amygdala, though has been long studied in the context of negative emotions, also plays a significant role in positive emotional representation ^39^ and reward learning ^40^. Earlier work indicates a role of amygdala projections in enhancing sensory processing of emotional stimuli ^41,42^. The excitatory connection from amygdala to visual cortex observed in our study provides empirical evidence for this notion and suggests that the amygdala may be able to increase attentional allocation toward stimuli associated with positively valenced experiences, thereby driving continued viewing.

Even when the content is less appealing, the activation of the amygdala persists, albeit at a reduced level. This finding further highlights the important role of amygdala in encoding valence and updating representations of value ^40,43^. Previous research has shown the amygdala contains detectors for both appetitive and aversive stimuli ^44^. Moreover, evidence from animal studies reveals an inhibitory relationship between these positive and negative encoding neurons ^45,46^. The amygdala’s putative capacity to represent a spectrum of subjective valences from pleasantness to unpleasantness may underpin its sustained engagement during continuous short-video viewing, with the activation level influenced by the interplay between populations of neurons encoding positive versus negative valence. And such encoding activity could plausibly impact choice selection and decision-making during media consumption ^47^. Further electrophysiological investigation is required to elucidate how the specific distribution and proportions of these neurons may affect amygdala activation and accompanying affective state in response to audiovisual stimuli.

The deactivation of System II during the viewing of preferred content further provides insights into the top-down mechanisms of video-watching behavior. The dlPFC is known for its involvement in executive functions such as working memory, cognitive flexibility, planning, inhibition, and abstract reasoning ^48^. The dACC is associated with conflict monitoring and adjusting control levels accordingly ^49^, while the preSMA is involved in changing movement plan and switching ^50^. The aIC is thought to integrate information from diverse functional systems and plays a crucial role in awareness, attention, and decision-making ^51^. It is believed that activation of these regions is essential for the implementation of specific cognitive processes ^52^, and the extent of their activation is also closely linked to task performance ^53^. In other word, if these brain regions displayed a negative BOLD signal (i.e., deactivated) during a cognitive task, then the functions they support are disabled or dysfunctional. Therefore, the observed deactivation of these regions during watching short-videos might contribute to the establishment of a “flow” state, a phenomenon where individuals are so absorbed in an activity that they lose track of time and their surroundings ^54^. Indeed, previous research has proposed that a flow state is associated with reduced frontal activity ^55,56^. Our finding confers preliminary empirical evidence for this theoretical postulation.

An intriguing phenomenon is that despite the overall deactivation of the control system, there exist individual variations in the level of inhibition, which correlate with individuals’ trait of mindfulness. That is, individuals with higher level of mindfulness exhibits less inhibition of the control system when consuming favorite video content, indicating that those individuals might still have a higher-level of awareness of present-moment experiences when facing tempting stimuli to prevent them from getting fully absorbed ^57^. Such association remains when facing less preferred content evoking behavioral adjustment, further suggesting that individuals with higher level of dispositional mindfulness might be more capable to effectively recruit their control system ^58^. Given that mindfulness training can improve awareness ^59^, we speculate that mindful practices may help to counteract the deactivation of System II seen during video watching, potentially promoting healthier video consumption habits with improved self-control abilities.

### The neural anatomic basis of the interplay between system I and II

The interplay between System I and System II during video watching provides a nuanced picture of how brain systems interact in this prevalent behavior. First, the inhibitory influences from the amygdala to prefrontal regions, as observed in our study, are supported by a substantial body of neuroanatomical and electrophysiological evidence. Specifically, the amygdala sends excitatory projections to the prefrontal cortex, but the overall influence of these inputs is predominantly inhibitory due to the preferential targeting of inhibitory interneurons within the PFC ^60,61^. Moreover, amygdala’s negative correlations with dlPFC, dACC, and aIC have been observed in a recent resting-state functional connectivity study ^62^, which is in line with aforementioned cellular-level findings. Second, dlPFC and dACC were found to have densely reciprocal connections with the amygdala ^63^. The feedback inhibition from dlPFC and dACC to amygdala align with earlier findings, as these regions are thought to downregulate the amygdala not only during explicit emotion regulation but also during cognitively demanding tasks ^64,65^. The context-dependent interaction of amygdala with prefrontal regions in video-watching behaviors might arise from the differentiated projections from the positive- and negative-encoding neurons in amygdala ^66^, but circuitry level evidence is required to test this conjecture.

The reciprocal inhibitory interaction between System I and System II contributes to a further refinement of the dual-system theory. Previous theory depicted their relationship as more of a see-saw battle ^22^, where the two systems compete in terms of activation strength, with the more strongly activated system determining final action ^23^. The inhibition of System II’s activity on System I has been supported, yet the impact of System I’s activity on System II remains unclear. Some studies put forth the hypothesis that heightened activation of System I might override System II, contributing to the loss of control to resist drugs ^67^. Our findings clearly pointed out that the hyperactivation of System I could suppress System II, going beyond the two-party competition on activation degree. Theoretically, if current affective stimuli keep System I highly involved, System II will remain suppressed. Consequently, individuals’ long-term goals cannot be represented in working memory, leading to a lack of prerequisites for regulating their behavior in the moment, even if it is inappropriate.

### Neuropsychological mechanisms of self-control during short-video viewing

The robust associations between personal trait self-control and brain metrics derived from video watching task and inner-state monitoring task, but not from the typical cognitive tasks, suggest a need to reconsider self-control related behaviors from a more ecological perspective. To this end and predicated on the dual-system theory, we proposed a Capture-Activate-Deactivate-Engage (CADE) model to account for the surge of short-video watching phenomena by taking the influence of powerful AI into account (Figure 6). This model elucidates the sequential progression of normal behaviors into problematic behaviors (e.g., unplanned binge-watching) through a series of four inter-locked stages as below:

- Capture - Attention is reflexively captured by highly salient video stimuli. This bottom-up capture of attention initiates preferential processing.
- Activate – The amygdala (System I), as a sensory and emotional processing hub, is activated to represent momentary emotional valence of the stimulus. If the amygdala’s activation is insufficient to satisfy one’s emotional expectation, the System II is recruited to disrupt the viewing process by scrolling to the next one. With such a voluntary selective iteration, the amygdala is eventually activated high enough to meet the psychologic satisfaction.
- Deactivate - The heightened activity of amygdala then inhibits prefrontal control regions (System II) through downstream effects on inhibitory interneurons. This suppresses goal representations and conflict monitoring functions subserved by these regions, with such a spectrum of individual difference that individuals with higher level of trait self-control are more capable to resist this suppression.
- Engage - With the cumulative effect of an activated System I and a deactivated System II, an immersed viewing state can be established progressively. While the amygdala activation accounts for surge of positive affective response that motivates users to consume more short-videos, the suppression of System II is responsible for a weakened awareness of one’s goals/plan, preventing ongoing experience being disrupted. As the AI algorithm is able to learn and recommend content based on each user’s preference, this Capture-Activate-Deactivate-Engage pathway is then constantly being reinforced, creating a closed-loop.

**Figure 6.**
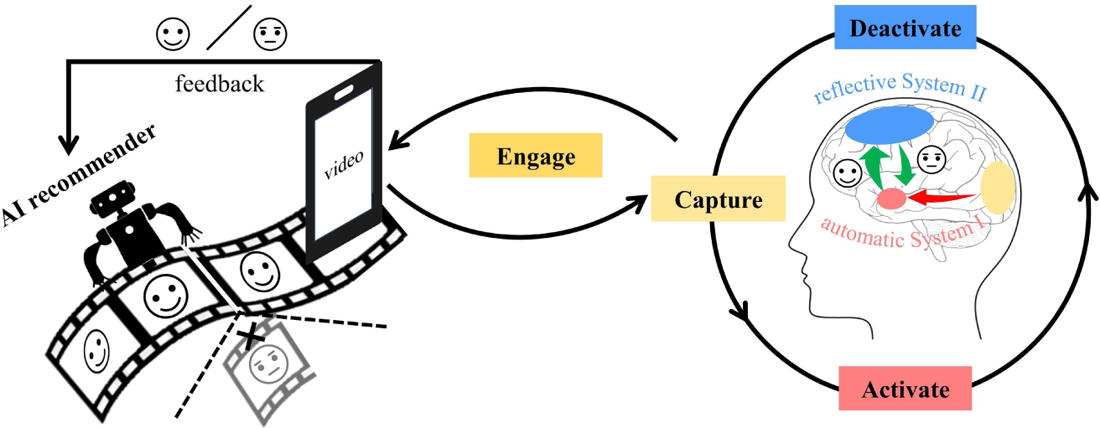
The Capture-Activate-Deactivate-Engage (CADE) Model. This model proposes four interlocked steps involving the dynamic interaction between the two systems during watching short-videos, with bias from recommendation algorithms designed to maximize user engagement. The process initiates with attentional **Capture** in face of salient video stimuli, which then **Activates** amygdala (System I) to construct emotional valence representation for subsequent behavioral choices. When the activation is high enough to meet user’s psychologic satisfaction, the amygdala **Deactivates** the prefrontal control regions (System II), subsequently weakening the representations of goals, plans, and self-awareness that rely on the neural activity of this system. Otherwise, an alternative video is explored until the psychologic satisfaction is achieved. With the cumulative effect of an activated System I and a deactivated System II, an immersed viewing state can be established progressively and users **Engage** in video watching. As AI algorithms are able to learn and recommend content based on each user’s preference, this CADE process is then continually being reinforced, creating a closed-loop.

This CADE model emphasizes the temporal dynamics involved in losing self-control during video watching. The initial bottom-up capture of attention combined with rapid deactivation of control regions allows an immersed state to swiftly engage, making voluntary restraint difficult.

There are several important implications of the CADE model. First, the CADE model highlights that individuals with less developed control system are likely more vulnerable to losing self-control during video watching. This predicts greater risk for younger populations like children and adolescents, whose prefrontal control systems are still maturing ^68,69^. Second, prolonged deactivation of control systems is another critical concern, as the persistent amygdala activation would activate its downstream interneurons in frontal regions, leading to the suppression of prefrontal control areas. The prefrontal inhibition, therefore, could be increased by the strengthened connection between amygdala and frontal interneuron through Hebbian learning principles ^70^. Aligning with this notion, excessive social media use is associated with impaired emotion regulation, like depression ^71^. Given the integrative role of prefrontal cortex in the human brain ^72^, careful examination of how uncontrolled viewing impacts self-control through the interactions between large-scaled brain network, is warranted. Third, the model highlights the potency of modern AI-driven tools for maximizing user engagement by continuously activating System I. If not properly monitored, algorithmic recommendations tailored based on user’s behavior might have the potential to influence democracy, privacy and mental health ^73,74^. Policy discussions around ethical constraints on engagement-maximizing algorithms are urgently needed to promote healthy technology use, particularly for younger users. Lastly, the positive association between self-control and activation of control brain regions during monitoring inner state suggests potential benefits of mindfulness training. The interoceptive awareness and non-judgemental acceptance ——two core elements fostered by mindfulness practice ^75^ —— might be an intrinsic source/power of breaking the above cycle ^76^. By enhancing metacognitive awareness of ongoing thoughts, emotions, and behaviors, mindfulness may empower individuals to exercise self-control capacities like conflict monitoring and top-down emotion regulation ^77,78^. Further research on how to optimize AI-powered algorithms to promote engagement in mindfulness practices may hold promise for counteracting unchecked technology use, offering fruitful new directions.

## Conclusion

Our findings shed new light on the neuropsychological mechanisms underlying self-control during video watching. First, we revealed the dynamics between System I and System II, with System I showing heightened activation and System II being suppressed during video watching, suggesting a lock-down of the control system especially when individuals were viewing their favorite content. Second, we found that the activation of System II regions during video watching and inner-state monitoring—but not during traditional cognitive tasks—correlated with self-control abilities, highlighting the crucial role of voluntary control over rule-based cognitive control in real-world self-control situations. This result bridges the gap between laboratory cognitive control tasks and real-world self-control over immediate gratification with a more ecologically valid paradigm. Third, the opposite effects of video-watching and inner-state mindful awareness on System II collectively underline the need for further exploration of the application of mindfulness practices in fostering self-control and healthier digital media use. In short, these findings may inform interventions for promoting healthier technology use and mitigating potential adverse effects of excessive screen time.

## Materials and Methods

### Participants

#### Sample one

Thirty-two healthy students were recruited to participate in this experiment at Zhejiang University. Five participants were excluded from analyses (four had too much head motion: maximum > 3mm, or more than 10% scrubbed volumes with frame-displacement (FD) ^79^ > 0.5; one disliked all short-videos). The final sample included 27 participants, with 17 males and 10 females, aged from 18 to 28 years (M = 22.56, SD = 2.28). All participants were right-handed, with normal vision, and reported no mental disease. Besides, they were experienced users of short-video apps and 23 out of 27 reported that they watched short-videos for more than 30 minutes per day. Participants in this sample completed a video-watching task and two cognitive tasks (Go/No-Go task and Dots task).

#### Sample two

We recruited another thirty-five students from Zhejiang university to perform an interoception task in this study (age between 19 and 28 years, M = 23.2, SD = 2.38, 15 male and 20 female). They were all healthy and reported no interoception-related disease. All participants were included for analyses. All participants signed written informed consent before attending the scanning. Each participant received monetary compensation for their time and travel. This study was approved by the Ethic Committee of Zhejiang University.

### Questionnaires used to measure trait self-control

#### Brief Self Control Scale

The Brief Self Control Scale (BSCS) developed by Tangney ^17^ has been validated to measure individual difference in general trait self-control. This study used a Chinese version of BSCS revised by Tan and Guo ^80^, which includes 19 items that assess five aspects: impulse control, work performance, healthy habits, entertainment restraint, and resisting temptation. All items are rated on a 5-point scale, ranging from 1 (strongly disagree) to 5 (strongly agree). The scores of five sub-scales were averaged to yield a total score. The higher scores signified greater levels of general trait self-control. The Cronbach’s alpha was 0.82 in our sample.

#### Barratt Impulsiveness Scale

The Barratt Impulsiveness Scale Version 11 (BIS-11) is commonly used to measure the impulsiveness with 30 items describing impulsive or non-impulsive behaviors and preferences ^81^. We used a well-validated Chinese version ^82^, and it had three factors: cognitive impulsiveness, motor impulsiveness, and lack of planning. Participants were required to rate items from 1 (never) to 5 (always). Higher scores indicated higher impulsive tendency, therefore, lower levels of trait self-control. The Cronbach’s alpha was 0.85 in the present study.

#### Five Factor Mindfulness Questionnaire

The Five Factor Mindfulness Questionnaire (FFMQ) developed by Baer ^83^, is a good instrument to assess individual’s trait level of mindfulness. It includes 39 items and taps five aspects of mindfulness: observing, describing, acting with awareness, nonjudging of inner experience, and nonreactivity to inner experience. Adding all the items’ score together yields a total score, and higher score reflects better mindfulness. The Chinese version used in the present study has been validated with acceptable psychometric properties ^84^. The Cronbach’s alpha was 0.79 in the present study.

#### Anxiety and Depression

The Brief Symptom Inventory 18 (BSI-18) ^85^, a shortened version of the 53-item Brief Symptom Inventory that was derived from Symptom Checklist-90, is a self-report checklist to measure psychopathological symptoms in the past week. It includes three dimensions: Somatization (6 items), Depression (6 items), and Anxiety (6 items). Participants need to rate 18 items based on a 5-point Likert-scale (0 for “not at all”, 1 for “a little bit”, 2 for “moderately”, 3 for “quite a bit”, 4 for “extremely”). In the present study, we only included Anxiety and Depression subscales and regarded them as potential control variables when calculating the relations between trait self-control and other brain measures. The Cronbach’s alpha of Anxiety and Depression was 0.79 and 0.88 in our data.

### Short-video watching task

A block design was employed in this task, consisting two 6-minute video-watching blocks and three 30-s rest blocks. A database with a total of 160 videos clips was built in advance. These videos were recorded from a popular short-video platform, using a newly registered account. An experimental operator with no history of using short-video apps is responsible for recording with a mobile device (Model: Xiaomi 9), during which she was instructed to record the short videos recommended by the platform without any personal bias. After the recording length reached one hour, the recording was halted. Then the experimenter trimmed the recorded videos and saved them for the final selection. The videos selected met two requirements: 1) the video content was of an entertaining nature and contained no violent, bloody, and political content; 2) each video was less than 2 minutes in length. Considering our samples were young university students, we included five categories of videos in our final database: single person showing action (e.g., singing, dancing, drawing, cooking, baby laughing and so on), multiple persons with interaction (e.g., sports, dance, family life, campus life), pets (mainly cats and dogs), game scenes, and natural scenery. The length of each short-video ranged from 7s to 104 s (Mean length=23.7s), and there are 130 videos with a duration of less than 30 seconds.

To simulate the real situation as much as possible when they were watching short videos in daily life, participants were merely instructed to relax during scanning and were allowed to switch to the next video clips at any time by pressing a button in their right hand. That meant each participant had greater autonomy in choosing which videos to watch and when to switch according to their own interests and preferences. Thus, videos can be naturally categorized as Like if participant kept watching them until the end, and as Dislike if participants switched before finishing a half. The videos that have being watched for more than a half but not completely were excluded from analyses.

The total number of short videos and their play order were predefined, but the actual amount of visited clips within each 6-minute block varied between participants. The average number of short videos that participants watched was 67.41 (SD = 13.88, ranging from 42 to 91). The number of videos were not significantly different between the Liked and Disliked categories (M = 35.21, SD = 20; M = 46.17, SD = 15.33; *t* = −1.688, *p* = 0.103). This task took 815 seconds.

### Go/No-Go task

An adapted Go/No-Go paradigm developed by Garavan ^86^ was used and the original stimuli (letter X and Y) were replaced by colored rectangles (red and green, size: 2cm×1cm) in the present study. Two conditions were included: 1) only colored rectangles presenting on the black background; 2) colored rectangles superimposed in the center of phased-scrambled pictures (13cm×6cm) presenting on the black background. Each condition involves both Go and No-Go trials. Participants were asked to press a button with the right thumb as quickly as possible when the colors of successively presenting rectangles were different (Go trials. e.g., red → green, or green → red), whereas to withhold the response if the colors did not change (No-Go trials. e.g., red → red, or green → green). Each rectangle was presented on the dark background for 800ms followed by a 200ms inter-stimulus interval. There were four blocks in this task and each block (80 trials) was followed by a 10s duration fixation cross. Among the total 320 trials, we set 44 No-Go trials therefore the ratio of No-Go trials to Go trials was 0.16:1. The reaction time on Go trials was calculated to measure motor execution and the rate of commission errors (i.e., failing to stop) on No-Go trials was used as an index for inhibition. The scan time for this task was 380 seconds.

### Dots task

The Dots task was first developed by Davidson et al ^87^, and we adapted a block-designed version used by Wang et al ^88^. In this task, three conditions were designed and participants were instructed to press button with their right or left thumb to make response to either a gray or stripped dot according to different rules. In the Congruent condition, only one type of dots (e.g., gray dots) were randomly presented on the left or right side of a central fixation, and participants needed to press spatially congruent buttons with corresponding thumbs. In the Incongruent condition, only the other type of dots (e.g., stripped dots) were appeared and participant should make side-incongruent responses. In the Mixed condition, both rules above were intermixed so it required participants to press buttons in response to two types of dots flexibly. Each condition consisted of three blocks, and each block lasted 26s, following a 12s fixation. The total trials in each condition were 36. To eliminate the fixed effect of certain rule, half of participants were asked to treat gray dots as congruent mark and treat stripped dots as incongruent mark; and the other half were trained with the reversed rules. For more details of task design, please see ^88^. In general, the incongruent condition has a greater demand on conflict monitoring and resolution than the congruent condition. The mixed condition has a greater demand on switching between different task sets compared to the incongruent condition. The mean reaction time and accuracy of correct trials in each condition was calculated to measure task performance. Considering the anticipatory response, we excluded trials with RT shorter than 200 ms before calculation. The duration of this task was 375 seconds.

### Interoception task

An inner-state tracking paradigm was used to assess the ability to monitor one’s present inner state^89^. There are two conditions in this task: 1) Heartbeat Counting (Heart), where participants were instructed to accurately detect and count their heartbeats over varied time intervals; and 2) Mental Counting (Count), wherein to control for the effects of mental counting on brain activity, participants were instructed to count numbers silently at a speed of 1 per second over the same intervals in the Heart condition, without attending to their heartbeat. A block design was employed, with both the Heart and Count conditions consisting of five blocks. The duration for heartbeat detection/counting varied across intervals of 20, 25, 35, 40, and 45 seconds, followed by a response window. Within each condition, the sequence of these durations was randomized. In each block of the Heart condition, a cue word “Heart” was presented for 3s. Subsequently, as the cue word was replaced by a fixation, participants began perceiving and counting their heartbeats. This fixation remained on the screen for the specified duration of the block, after which it was replaced by the number “55”. Participants reported their heartbeats by pressing 4 buttons to adjust the two “5” digits by an increase or decrease of 1. In the Count condition, the cue word was changed to “Count” and participants were instructed to count numbers. The Heart and Count conditions were presented alternatively. A fingertip blood pressure monitor (compatible with the Siemens 3.0-T scanner) was put on the participant’s left index fingers to record their heart rate during the task to verify that the participants follow the instruction.

### Experimental procedure

For both samples, participants were given a tutor about the whole experimental procedure and signed the consent form upon arrivals. In the first sample, participants completed questionnaires first. Then, a training session was provided to help them understand the rules of each task with practice. Specifically, for the short-video watching task, participants were instructed to choose the videos to watch using the button on their right hand. For Go/No-Go and Dots tasks, participants performed practices and only with accuracy greater 85% could they move forward to formal testing. Another practice chance would be provided if participants failed. If the accuracy rate in the second practice session was still below 85%, we inquired about the participant’s state, reiterated the task rules, and asked them to practice for a third time. In our sample, there was only one participant who needed three practice sessions for the Go/No-Go task. In the scanner, participants underwent an 8-minute resting-state scan and then the short-video task, Go/No-Go task, and Dots task in order. For participants in the second sample, they practiced on the Heart and Count conditions and learn to use the four buttons for report outside the scanner and underwent a resting-state scan and then the interoception task inside the scanner. The questionnaires were completed after scanning for the second sample. All the resting-state data were not analyzed in the present work.

All stimulus presentation and response acquisition were performed within the E-Prime 3.0 environment (Psychology Software Tools, Inc., Pittsburgh, PA). Stimuli were presented through an MRI compatible screen with 720× 1280 pixels resolution. Foam padding were used to limit head movement, and noise-cancelling headphones were provided to reduce scanner noise for participants.

### Image data acquisition

Brain imaging data were collected in a Siemens 3.0-T scanner (MAGNETOM Prisma, Siemens Healthcare Erlangen, Germany) with a 20-channel coil. Structural images were acquired with a T1-weighted magnetization prepared rapid gradient echo sequence (TR = 2300 ms, TE = 2.32 ms, voxel size = 0.90 × 0.90 × 0.90 mm^3^, voxel matrix = 256 × 256, flip angle = 8°, field of view = 240 × 240). Functional images were collected using a gradient echo planar imaging sequence with multi-band acceleration (factor = 4, TR = 1000 ms, TE = 34 ms, voxel size = 2.50 × 2.50 × 2.50 mm^3^, voxel matrix = 92 × 92, flip angle = 50 °, field of view = 230 mm, slices number = 52).

### Image preprocessing

The fMRI data were pre-processed using AFNI ^90^ with common routines including slice timing, head motion correction, normalization, and smoothing (FWHM= 5mm). The segmentation was conducted to extract brains using SPM12 (https://www.fil.ion.ucl.ac.uk/spm/). Structural and functional images were normalized to the MNI space using ANTs (http://stnava.github.io/ANTs/).

### First level general linear modelling (GLM)

The GLM was conducted to estimate the brain response to each task condition, using the command 3dDeconvolve in AFNI.

- For the video-watching task, three block regressors, namely Like, Dislike, and ‘Unclassified’, were constructed to characterize the brain responses to videos being viewed to the end, being switched before half, and being switched after half, respectively. The onset times were the times when the video appeared on the screen and the durations were the lengths of the video being viewed. As the participants made button presses under Dislike and Unclassified conditions, two event-related regressors were constructed to account for the fMRI signal changes associated with motor responses in the two conditions. Brain responses to Dislike and Like conditions during the viewing period and their contrast were of the primary interest for this task.
- For Go/No-Go task, four event-related regressors were created for successful Go trials, successful NoGo trials, failed Go trials, and failed NoGo trials. The main contrasts of interest were the brain responses to the correct NoGo (abbr. as NoGo), the correct Go (abbr. as Go) conditions and their difference.
- For the Dots task, three block-wise regressors were created for the Congruent, Incongruent, and Mixed conditions.
- For the interoception task, two block-wise regressors were constructed for the Heart and Count conditions when participants making heartbeat or number counting, respectively. Four event-related regressors were created to capture the transient response (onset of each operation cue) at the start and end of each block, separately for the two conditions. The two block-wise regressors and their contrast were of the primary interest in the present work.

In addition to the task related regressors, regressors of nuisance, including 6 head motion parameters, signal drifts (automatically determined with the option-polort A in 3dDeconvolve command) in all the tasks. As signals from CSF and WM have been shown to contain physiology noise (e.g., heart rate and respiration) ^91^, the first five components of CSF and WM were included as confounds for all the tasks except for the interoception task as this task required heartbeat perception.

### Whole brain voxel-wise analyses at the group level for the video task

For the video-watching task, one sample t-tests were conducted to identify brain activation associated with each condition, and a paired two-sample t-test was used to compare the differences between conditions of Like and Dislike. Multiple comparison correction was accomplished using the 3dClustSim mixed-model autocorrelation function (ACF) in AFNI. A corrected significance level of *p* < .05 could be achieved with a minimum cluster size of 30 voxels when the threshold was set at *p* < .001.

### ROI analyses on System I and II

To further characterize the neural activities of key brain regions modulated by video task in different context, a region of interest (ROI) approach was adopted to analyze the two cognitive control tasks (i.e., Go/No-Go and Dots) the interoception task. Based on our hypothesis on System I and II, we focused on the activity and interaction of two systems. Therefore, the amygdala regions showing activation difference between Like and Dislike condition were selected to represent System Ⅰ. Similarly, the dlPFC, dACC, preSMA, and aIC were selected as four vital regions in System Ⅱ based on the activation difference between the two conditions in the video task, with a reference to a meta-analysis result with the term of “control” in Neurosynth (supplementary Figure S3, https://neurosynth.org/analyses/terms/control/).

More specifically, a binary mask was saved from the contrast activation map (Like > Dislike, corrected p < 0.05). The saved mask was: 1) multiplied with the Brodmann’s area 9 and 46 to create the mask for dlPFC ROI; 2) multiplied with the Automated anatomical labelling atlas (AAL 90) ^92^ masks for cingulate (labelled 31 and 32) to create the dACC ROI; 4) multiplied with AAL masks labelled 33 and 34 to create the preSMA ROI. The masks of amygdala and aIC from task activation map were directly taken as corresponding ROIs as they were well constrained within the AAL masks for amygdala and insula. Mean beta values of these ROIs were extracted from each condition for each of the three tasks for each participant, and were used in below statistical analyses.

One-sample t-tests were used to examine each ROI’s activation under each condition across tasks. Paired-sample t-test was conducted to compare the activation difference between Go and No-Go trials for the Go/No-go task. One-way repeated ANOVAs were conducted to compare the beta values among the three conditions in the Dots task. Correlations between the beta values of the five ROIs and self-reported self-control scores from BSCS, BIS-11, and FFMQ were examined, controlling for age, gender, anxiety and depression scores. Group level mean value and individual data points were used for plotting bar graph (Figure 3). Statistical analyses of behavioral and imaging data were performed using SPSS (version 22.0, https://www.ibm.com/analytics/spss-statistics-software) and JASP (version 0.18.0, https://jasp-stats.org/).

### Dynamic Causal Modelling (DCM)

DCM can be used to delineate neuronal dynamics through bilinear approximations ^37^, usually in terms of effective connectivity between regions of interest. Therefore, we implemented the DCM, a toolbox implemented in SPM 12 (https://www.fil.ion.ucl.ac.uk/spm/software/), to characterize the interaction between the two systems, using a deterministic, one-state model for fMRI. This model depicts the derivative of neural state at any given time as a function of the current state (z), the experimental input (u), and parameters that determine the strength of connections within and between brain regions:

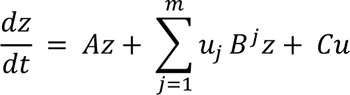

where the matrix A represents endogenous connections that are not affected by external input, including inhibitory self-connection and between-region connections; the matrix B is the modulatory effect exerted by experimental manipulations on the connectivity; and the matrix C stands for the direct effect on each region due to driving inputs.

#### VOI definition and timeseries extraction

Following previous work ^93^ and the DCM guide ^94^, five regions in the right hemisphere of the brain were selected as volumes of interest (VOI) from the video-watching task for subsequent DCM analysis, with a two-step selection procedure detailed below. The first step was to define VOI at the group level. Specifically, group level peak voxels were identified for five cortical regions as below. 1) the group-level coordinate of peak voxel in visual cortex (MNI: x = 14.5, y = −100, z = 1) from the main effect of task (watching video clips vs. rest) was identified as the information input node of the DCM network. 2) the dACC (MNI: x = 4.5, y = 22.5, z = 31), 3) preSMA (MNI: x = 2, y = 17.5, z = 46), 4) aIC (MNI: x = 32, y = 22.5, z = 1), and 5) dlPFC (MNI: x = 39.5, y = 42.5, z = 28.5) were identified as the peak voxel from contrast of Like > Dislike. Considering the complex anatomy and small volume of amygdala (relative to cortex), an amygdala mask from meta-analysis (https://neurosynth.org/analyses/terms/amygdala/) (setting z above 30) was used for individualized signal extraction in the second step detailed below. The second step was to extract signals from individualized VOI, constrained by the group level VOI. Specifically, for every subject, a peak voxel was first searched within a distance of 8 mm from the group peak coordinate, and then voxels within a 4-mm-radius sphere centered at the peak coordinate were included into the final VOI used in DCM if the voxel showed a threshold of p<0.05 for the effect of interest. The threshold was allowed to be even more liberal if no voxel was found ^94^. The effects of no interest (nuisance regressors), including the motor responses and movements, were regressed out by adjusting a F-contrast, and the first principal eigen-variate time series of each ROI was extracted.

#### Specification of the model

First, a full model was set for each participant. The visual cortex was assumed to have bidirectional connection with amygdala ^95^, and the other five regions were all bidirectionally connected. The “task”, including all visual stimuli during watching video clips, was set as a single driving input on visual cortex only. Like and Dislike both functioned as modulatory input on all possible connectivity specified in the matrix A, except for the intrinsic self-inhibition of visual area. Without applying the mean-centred option to experimental input, the A matrix here represented the connectivity of baseline (fixation).

#### Model estimation

Then, this model was inverted using Variational Laplace to evaluate the quality of the model and obtain a probability density over parameters. After completion of this estimation process, we checked each subject’s explained variance and identified five of them with relatively poor explained variance (below 10%). We excluded them in subsequent group-level analysis based on the previous technique paper regarding the application of DCM ^94,96^.

#### Group-level analysis

Next, Parametrical Empirical Bayes (PEB) was implemented to quantify the group mean of connection strength and the differences across subjects ^97^. The covariate of interest (i.e., the group mean) was included in the first column of design matrix X, and anxiety, depression, age, and gender were also added as nuisance covariates. All of these five regressors were mean-centered. Without a prior knowledge of how would the connectivity be modulated by Like and Dislike input, the auto-search Bayesian model reduction (BMR) routine was applied to select the best model. Three separate PEB (A, B, and C) analyses were carried out, using greedy search to iteratively prune parameters that did not contribute to the model evidence from the full PEB model. Then parameters were further averaged using the Bayesian model averaging (BMA), and only parameters with a posterior probability > 95% were reported.

## Acknowledgment

This research was supported by the STI 2030 Major Projects (No. 2021ZD0200409); National Natural Science Foundation of China (No. 81971245, 62077042); the Zhejiang Province “Qianjiang Talent Program”, and the MOE Frontiers Science Center for Brain Science & Brain-Machine Integration, Zhejiang University.

## Data availability

Due to ongoing utilization of the raw fMRI data in other analyses, full disclosure of it is currently not feasible. However, it is available upon request by contacting the corresponding author (email address).

## Supplementary Materials

### 1. The brain regions showing significant difference in activation between Like and Dislike conditions in video-watching task

**Table S1.**
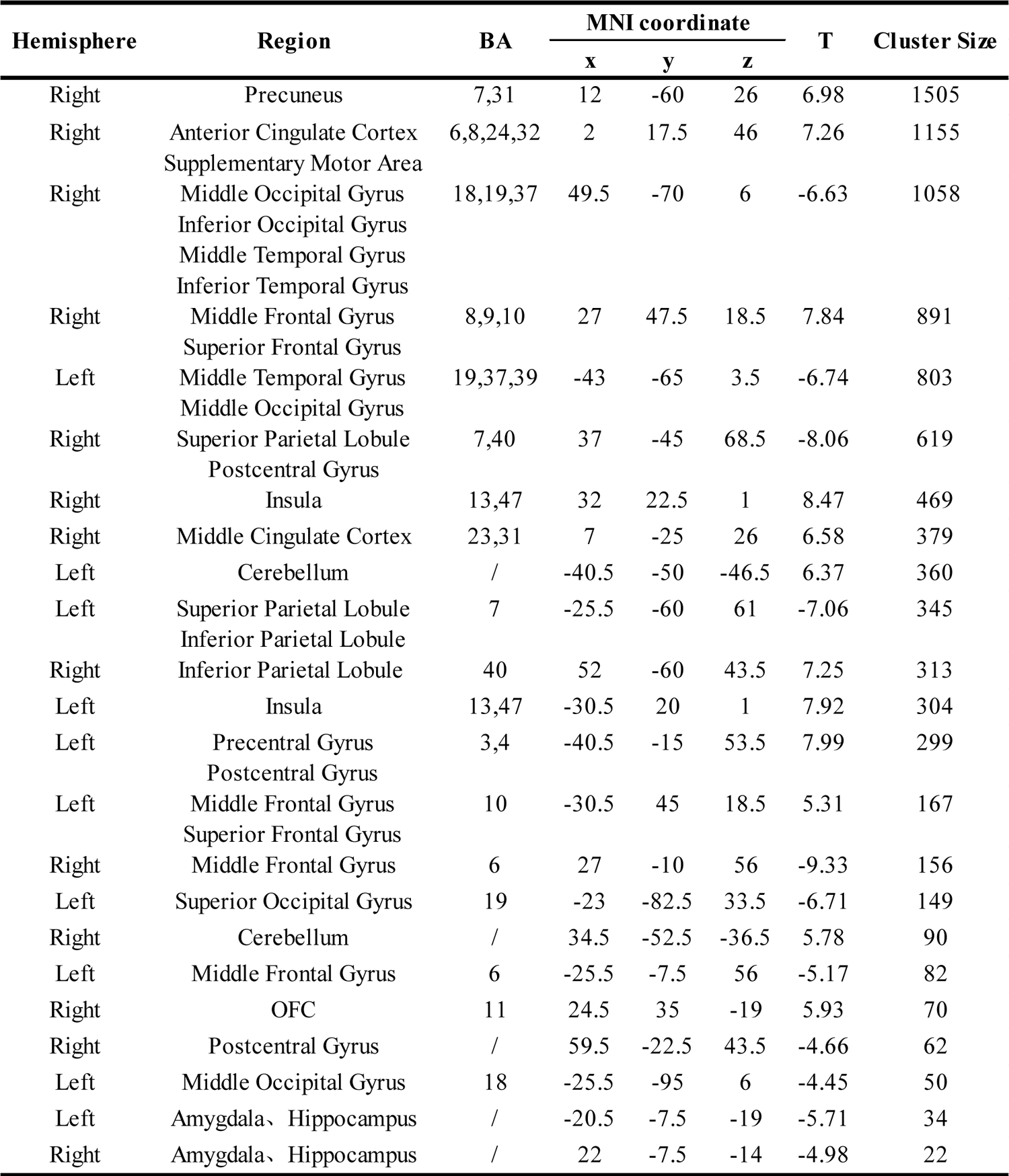

### 2. The LOOCV result of correlation between trait self-control and brain activation

#### 2.1 Aim

To examine the robustness of the relationship between brain activities and self-reported measurement on self-control, validation was performed by using a leave-one-out analysis via MATLAB code (shared by Grigori Yourganov. https://github.com/grigori-yourganov/leave_one_out).

#### 2.2 Methods

Linear regression was used in present analysis, using the beta value of dlPFC, dACC, preSMA and aIC extracted from the Dislike condition as predictors (together with anxiety, depression, gender, and age) to predict the BSCS, BIS-11 and FFMQ, respectively. For every iteration, one observation was removed and its outcome was predicted by an estimated model using all other observations. After finishing iteration, the previous excluded observation was put back into the origin data set and the next observation was removed to perform a new iteration. The above procedure would be repeated until all observations have been selected. We calculated the Pearson’s correlation between actual and predicted scores of BSCS, BIS-11 and FFMQ, and regarded it as a measure of generalization of current result.

#### 2.3 Result

As shown in the Figure S1, the trait self-control (BSCS, BIS-11, and FFMQ) predicted by the LOOCV model has a good fitting with their actual values. Their correlation coefficients are moderate and all achieve significance (*p* < .05), except for BSCS predicted by the activity of preSMA under the dislike condition, which has a marginal significance (*p* = .06).

**Figure S1.**
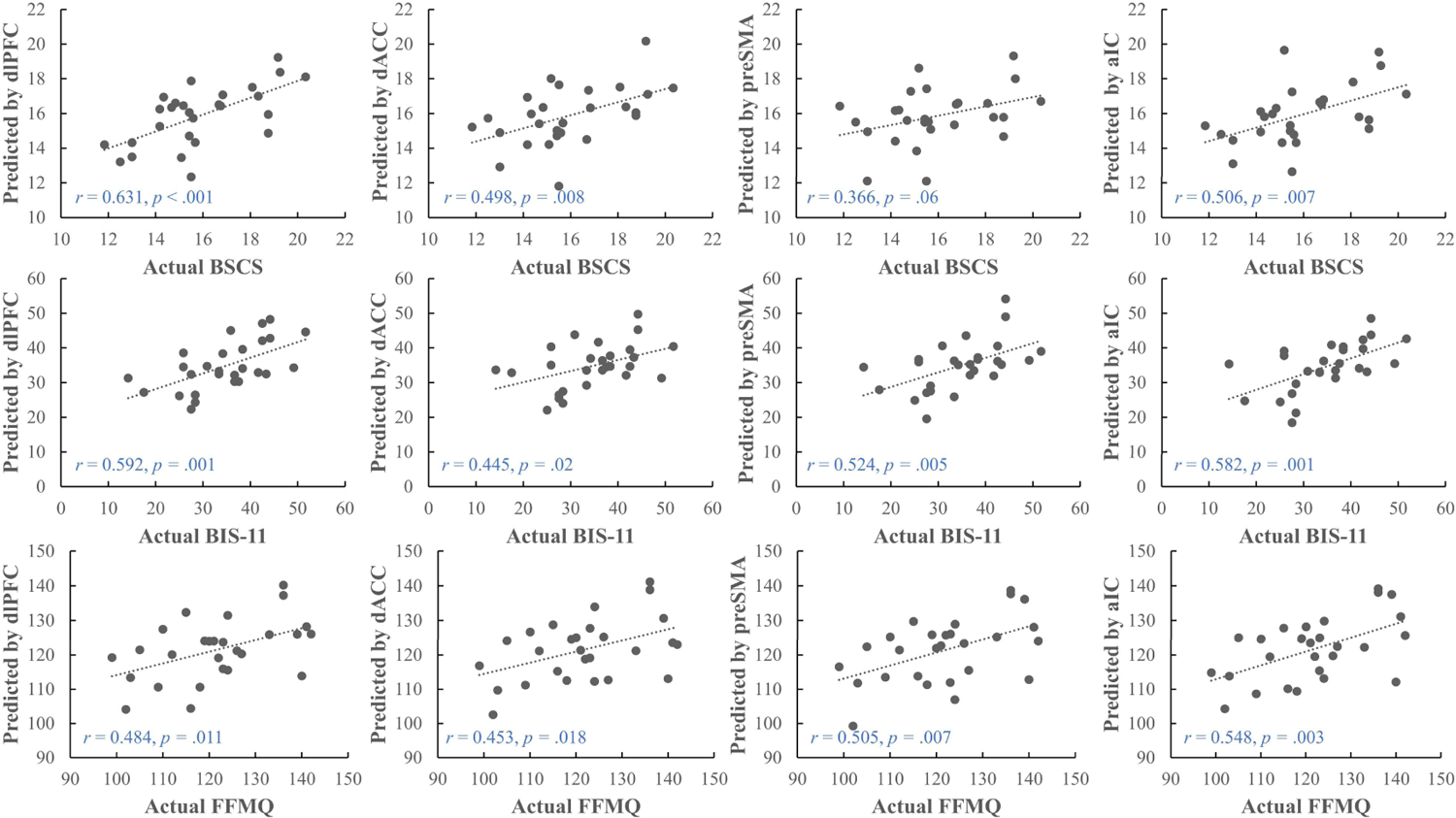
The scatterplots of leave-one-out validation result. The top row shows the Pearson’s correlation between the actual self-control scores (BSCS) and their estimated value predicted by the regression coefficient (beta) of dlPFC (left), dACC (middle), preSMA (middle) and aIC (right) under the Dislike condition. The middle row represents the predicted results for impulsivity (BIS-11). And the bottom row depicts the results for mindfulness (FFMQ).

### 3. The full model for DCM and the PEB results

**Figure S2.**
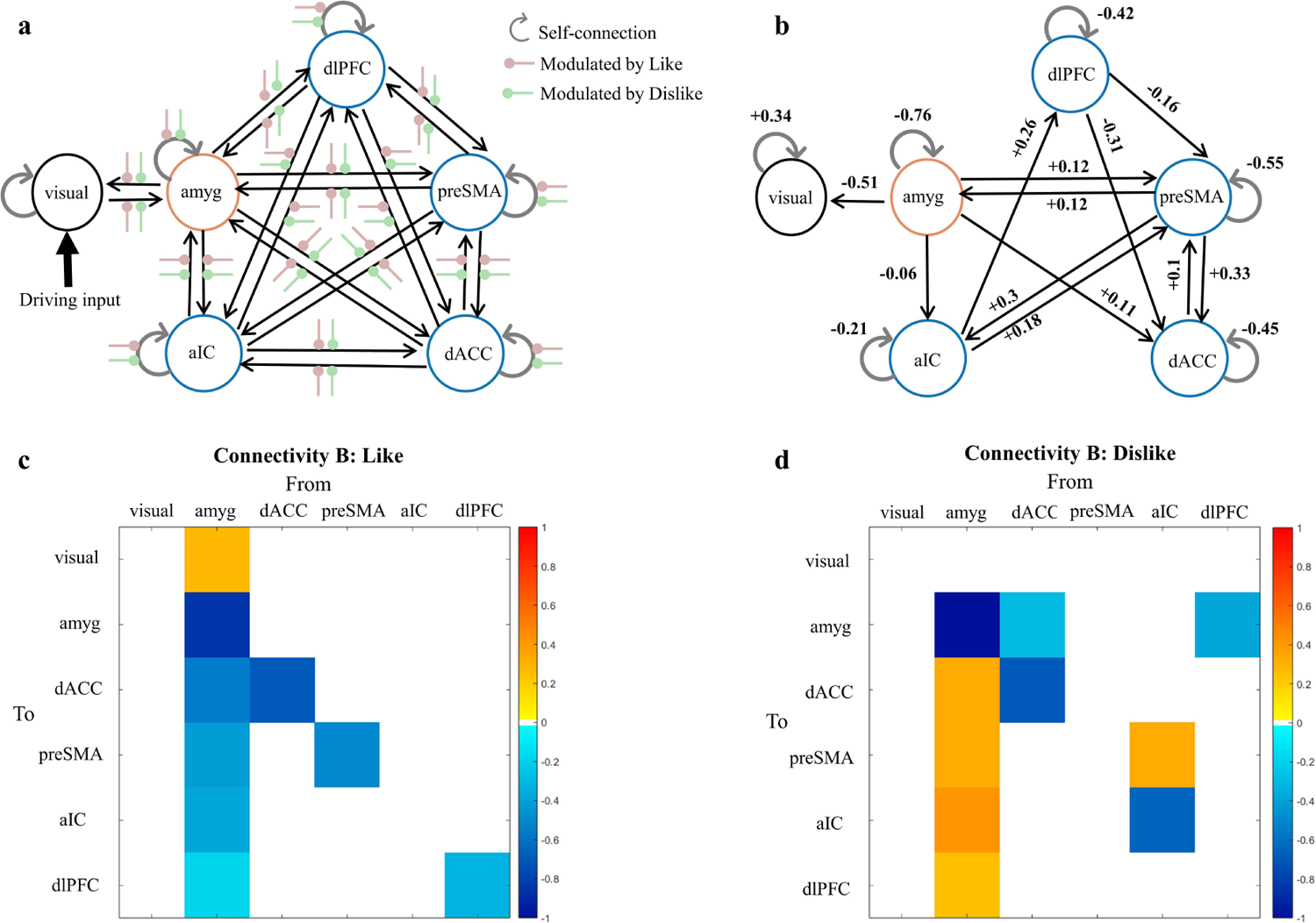
The schematic diagram of the full model for DCM (a) and the group-level PEB results (b, c, d). The parameters in matrix “A” represent baseline effective connectivity (b) and the two matrices “B” show modulatory parameters by Like (c) and Dislike (d) conditions. Only parameters with a posterior probability above 95 % were shown. Parameters on the leading diagonal of matrices are self-connection (unitless) and the off-diagonal parameters are between-region connection (rate of change, in units of hertz). Positive numbers (warm colors) indicate excitatory connection and negative ones (cool colors) mean inhibitory connection. Abbreviations: amyg, amygdala; dACC, dorsal anterior cingulate cortex; preSMA, pre-supplementary motor area; aIC, anterior insular cortex; dlPFC, dorsolateral prefrontal cortex.

### 4. Descriptive information on task performance and its correlation with trait self-control

#### 4.1 Aim

As self-control is a multidimensional concept, we also examined the relationship between the behavioral performance of the three tasks and three measurements of trait self-control.

#### 4.2 Method

In the video watching task, we focused on three indicators, including the total number of short videos each participant watched (Video num in Table S2), the proportion of videos being watched to end (Like rate in Table S2) and that for these being switched (Dislike rate in Table S2).

For the Go/No-Go task, the mean reaction time of successful Go trials and the rate of error of NoGo trials (Commissions errors in Table S2) were used to characterize task performance.

For the Dots task, the performance was indicated by the average response time and accuracy across the three conditions (i.e., Congruent, Incongruent, and Mixed in in Table S2).

The descriptive statistic results of these task measures and questionnaires were listed in the table S2. After controlling for age, gender, anxiety, and depression, we calculated the partial correlation between task indicators and self-report self-control.

#### 4.3 Result

As shown in Table S2, the BSCS score showed significant correlation with BIS-11 but not FFMQ, but the latter two correlated with each other, indicating the three questionnaires, while all relate to trait self-control, may capture some distinct aspects of trait self-control.

Regarding the relationships between trait self-control measures and behavioral measures from experimental tasks, most correlations were not significant. Only the average response time under the Mixed condition of the Dots task was significantly positively correlated with BSCS, and significantly negatively correlated with BIS-11, but not significantly correlated with FFMQ.

**Figure S3.**
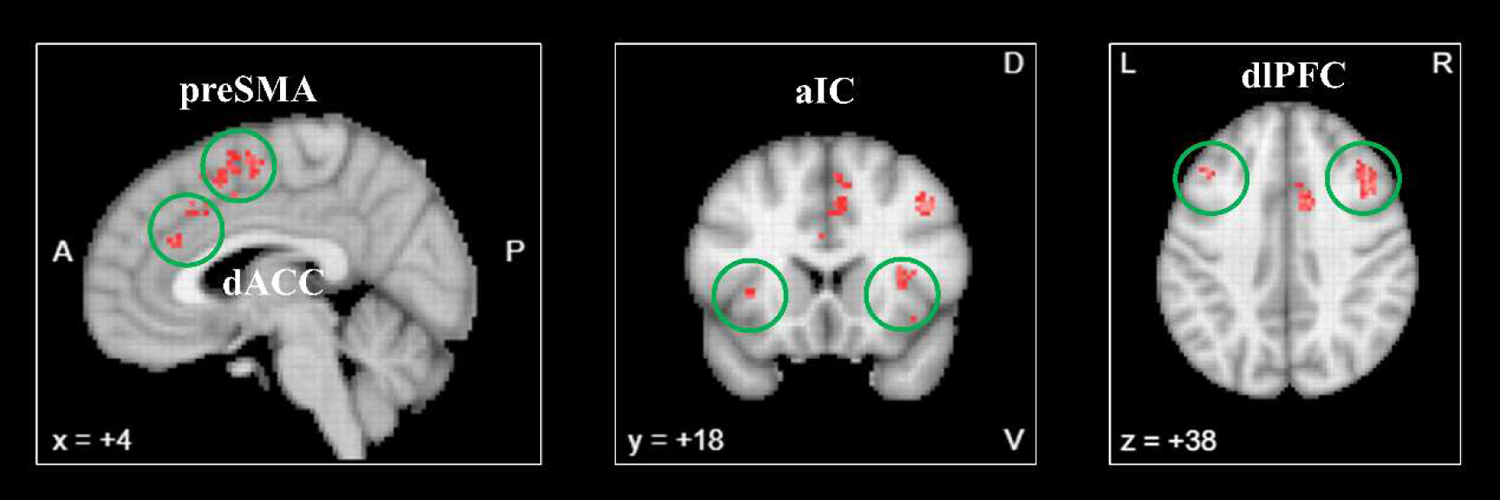
The activation map of “control” generated from meta-analysis of 3796 studies. Here we showed the association test map, from which we selected four brain regions (highlighted by the green circles) to represent the control system (i.e., System II). These regions included dorsal anterior cingulate cortex (dACC), pre-supplementary motor area (preSMA), anterior insula cortex (aIC), and dorsolateral prefrontal cortex (dlPFC). Picture source: https://neurosynth.org/analyses/terms/control/

### 5. Supportive evidence for System II ROI selections based on meta-analysis

**Table S2.**
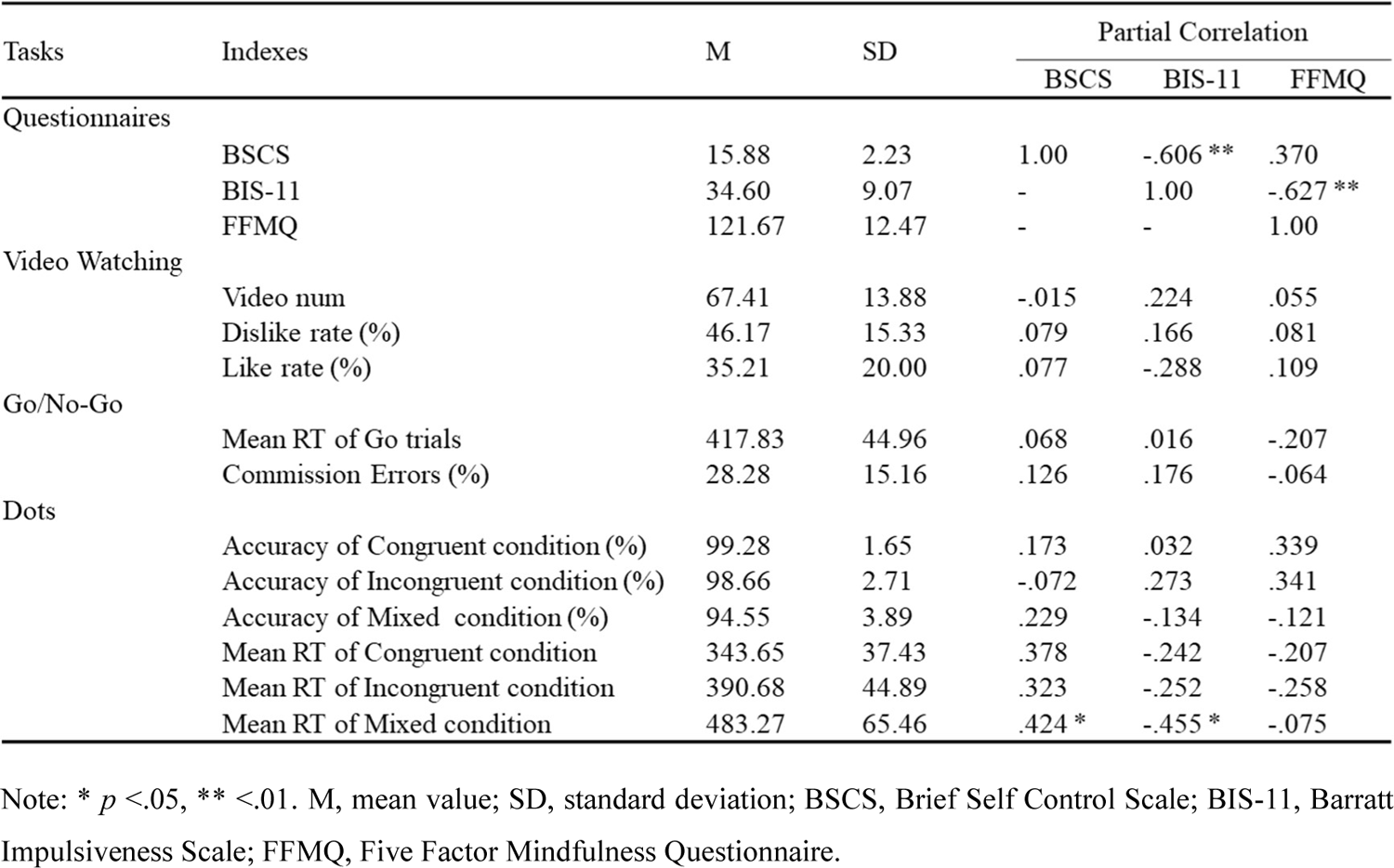
Statistical information about questionnaires and behavioral indexes.

| Tasks | Indexes | M | SD | Partial Correlation |  |  |
| --- | --- | --- | --- | --- | --- | --- |
|  |  |  |  | BSCS | BIS-11 | FFMQ |
| Questionnaires |  |  |  |  |  |  |
|  | BSCS | 15.88 | 2.23 | 1.00 | -.606 ** | .370 |
|  | BIS-11 | 34.60 | 9.07 | - | 1.00 | -.627 ** |
|  | FFMQ | 121.67 | 12.47 | - | - | 1.00 |
| Video Watching |  |  |  |  |  |  |
|  | Video num | 67.41 | 13.88 | -.015 | .224 | .055 |
|  | Dislike rate (%) | 46.17 | 15.33 | .079 | .166 | .081 |
|  | Like rate (%) | 35.21 | 20.00 | .077 | -.288 | .109 |
| Go/No-Go |  |  |  |  |  |  |
|  | Mean RT of Go trials | 417.83 | 44.96 | .068 | .016 | -.207 |
|  | Commission Errors (%) | 28.28 | 15.16 | .126 | .176 | -.064 |
| Dots |  |  |  |  |  |  |
|  | Accuracy of Congruent condition (%) | 99.28 | 1.65 | .173 | .032 | .339 |
|  | Accuracy of Incongruent condition (%) | 98.66 | 2.71 | -.072 | .273 | .341 |
|  | Accuracy of Mixed condition (%) | 94.55 | 3.89 | .229 | -.134 | -.121 |
|  | Mean RT of Congruent condition | 343.65 | 37.43 | .378 | -.242 | -.207 |
|  | Mean RT of Incongruent condition | 390.68 | 44.89 | .323 | -.252 | -.258 |
|  | Mean RT of Mixed condition | 483.27 | 65.46 | .424 * | -.455 * | -.075 |

Note: \* $p < .05$ , \*\* $p < .01$ . M, mean value; SD, standard deviation; BSCS, Brief Self Control Scale; BIS-11, Barratt Impulsiveness Scale; FFMQ, Five Factor Mindfulness Questionnaire.

